# Efficacy and safety of ensitrelvir in patients with mild-to-moderate COVID-19: the phase 2b part of a randomized, placebo-controlled, phase 2/3 study

**DOI:** 10.1101/2022.06.22.22276792

**Authors:** Hiroshi Mukae, Hiroshi Yotsuyanagi, Norio Ohmagari, Yohei Doi, Hiroki Sakaguchi, Takuhiro Sonoyama, Genki Ichihashi, Takao Sanaki, Keiko Baba, Yuko Tsuge, Takeki Uehara

## Abstract

This phase 2b part of a randomized phase 2/3 study assessed the efficacy and safety of ensitrelvir for mild-to-moderate coronavirus disease 2019 (COVID-19). Patients were randomized (1:1:1) to orally receive ensitrelvir fumaric acid 125 mg (375 mg on day 1; *n*=140) or 250 mg (750 mg on day 1; *n*=140) or placebo (*n*=141) once daily for 5 days. Compared with placebo, the change from baseline in severe acute respiratory syndrome coronavirus 2 titer (measured as log_10_ 50% tissue-culture infectious dose) on day 4 was significantly greater with ensitrelvir 125 mg and 250 mg (differences from placebo: −0.41, *P*<0.0001 for both). The total score of predefined 12 COVID-19 symptoms showed an improving trend with ensitrelvir treatment without a significant intergroup difference. Most adverse events were mild in severity. Ensitrelvir treatment demonstrated a favorable antiviral efficacy and potential clinical benefit with an acceptable safety profile. (Japan Registry of Clinical Trials identifier: jRCT2031210350)

## Main

Coronavirus disease 2019 (COVID-19), caused by infection with severe acute respiratory syndrome coronavirus 2 (SARS-CoV-2), has been declared a global pandemic by the World Health Organization (WHO) (https://www.who.int/emergencies/diseases/novel-coronavirus-2019). As of May 25, 2022, more than 520 million confirmed cases of COVID-19 have been reported worldwide, with over 6 million COVID-19–associated deaths (https://covid19.who.int/). To combat the COVID-19 pandemic, several vaccines against SARS-CoV-2 infection have been developed and approved for clinical use^1–4^, and more than 11 billion vaccine doses have been administered to the worldwide population (https://covid19.who.int/). However, humoral immune response against SARS-CoV-2, as measured by neutralizing antibody titers, decreases substantially with time after vaccination^5^, potentially leading to postvaccination infections^6^. Therefore, in addition to vaccines, effective antiviral agents are needed for the management of the COVID-19 pandemic.

Gene mutations in the SARS-CoV-2 genome have resulted in the emergence of various variants, of which those posing an increased risk to global health are referred to as variants of concern (VOCs). The SARS-CoV-2 Omicron (B.1.1.529) variant was first detected in South Africa in November 2021, quickly spread worldwide, and was designated as a VOC by the WHO (https://www.who.int/news/item/26-11-2021-classification-of-omicron-(b.1.1.529)-sars-cov-2-variant-of-concern). By the end of January 2022, >99% of SARS-CoV-2 cases reported in the United States (US) had been infected with the Omicron variant^7^. This variant is characterized by more than 30 mutations located in its spike protein, of which 15 are focused on the receptor-binding domain^8^. Mutations in the receptor-binding domain of the spike protein result in high binding affinity with human angiotensin-converting enzyme 2, increased viral invasion into host cells, and a high transmission rate associated with this VOC^9^. The disease caused by infection with the Omicron variant is considered to be less severe than that caused by previous variants^10–12^, but the high transmissibility and infectivity of this variant may pose an additional threat to global health security. Several antiviral treatment options for patients with COVID-19 at risk of severe disease, including molnupiravir^13^, nirmatrelvir in combination with a pharmacokinetic booster ritonavir^14^, sotrovimab^15^, and casirivimab/imdevimab^16^, have demonstrated efficacy in reducing the risk of hospitalization, death, or disease progression. However, in view of the public burden of the transmission of infectious viruses and disease characteristics of COVID-19, as well as to draw maximum benefits from early pharmacological treatment initiation (https://www.kansensho.or.jp/uploads/files/topics/2019ncov/covid19_drug_220218.pdf; website in Japanese), additional oral antiviral agents that can be administered to patients with mild-to-moderate COVID-19 regardless of the risk of severe disease are needed.

Ensitrelvir fumaric acid (S-217622; hereafter, ensitrelvir), a novel oral SARS-CoV-2 3C-like protease inhibitor, was discovered through joint research by Hokkaido University and Shionogi & Co., Ltd^17^. Ensitrelvir has shown antiviral efficacy in both *in vitro* and *in vivo* animal studies against different SARS-CoV-2 variants, including the Omicron variant^17–19^. Additionally, in a phase 1 study of ensitrelvir (Japan Registry of Clinical Trials identifier: jRCT2031210202), the once-daily oral dose was well tolerated and demonstrated a favorable pharmacokinetic profile (R. Shimizu, et al., unpublished data). A multicenter, randomized, double-blind, placebo-controlled, phase 2/3 study is also underway to assess the efficacy, safety, and pharmacokinetics of ensitrelvir administered as a 5-day oral regimen. In its phase 2a part, although the sample size was small, ensitrelvir treatment led to a reduction in the SARS-CoV-2 viral titer and viral RNA versus placebo with an acceptable safety profile^20^. Herein, we present the antiviral and clinical efficacy and safety of ensitrelvir derived from the phase 2b part of this phase 2/3 study.

## Results

### Patient disposition

This phase 2b, dose-finding part of the study (Japan Registry of Clinical Trials identifier: jRCT2031210350) was conducted from January 2 to February 9, 2022, at 87 institutions in Japan and South Korea (Supplementary Table 1). Of the 437 patients who provided informed consent, 9 were excluded prior to randomization due to screening failure. Among the 428 patients in total, 142, 143, and 143 were randomized to receive ensitrelvir 125 mg (375 mg on day 1), ensitrelvir 250 mg (750 mg on day 1), or placebo, respectively, until day 5; 140, 140, and 141 patients, respectively, were included in the safety analysis population. A total of 134, 136, and 139 patients in the ensitrelvir 125 mg, ensitrelvir 250 mg, and placebo groups, respectively, completed the study. After the exclusion of patients with undetectable SARS-CoV-2 titer at baseline or those who provided incomplete informed consent (ensitrelvir 125 mg, 28; ensitrelvir 250 mg, 27; and placebo, 32; some patients were excluded from the analysis populations due to more than one reason), 341 patients (ensitrelvir 125 mg, 114; ensitrelvir 250 mg, 116; and placebo, 111) were included in the intention-to-treat (ITT) population (Fig. 1). All analyses were performed in the planned treatment groups. The ITT population was used for all efficacy analyses, and the safety analysis population was used for all safety analyses.

**Fig. 1:**
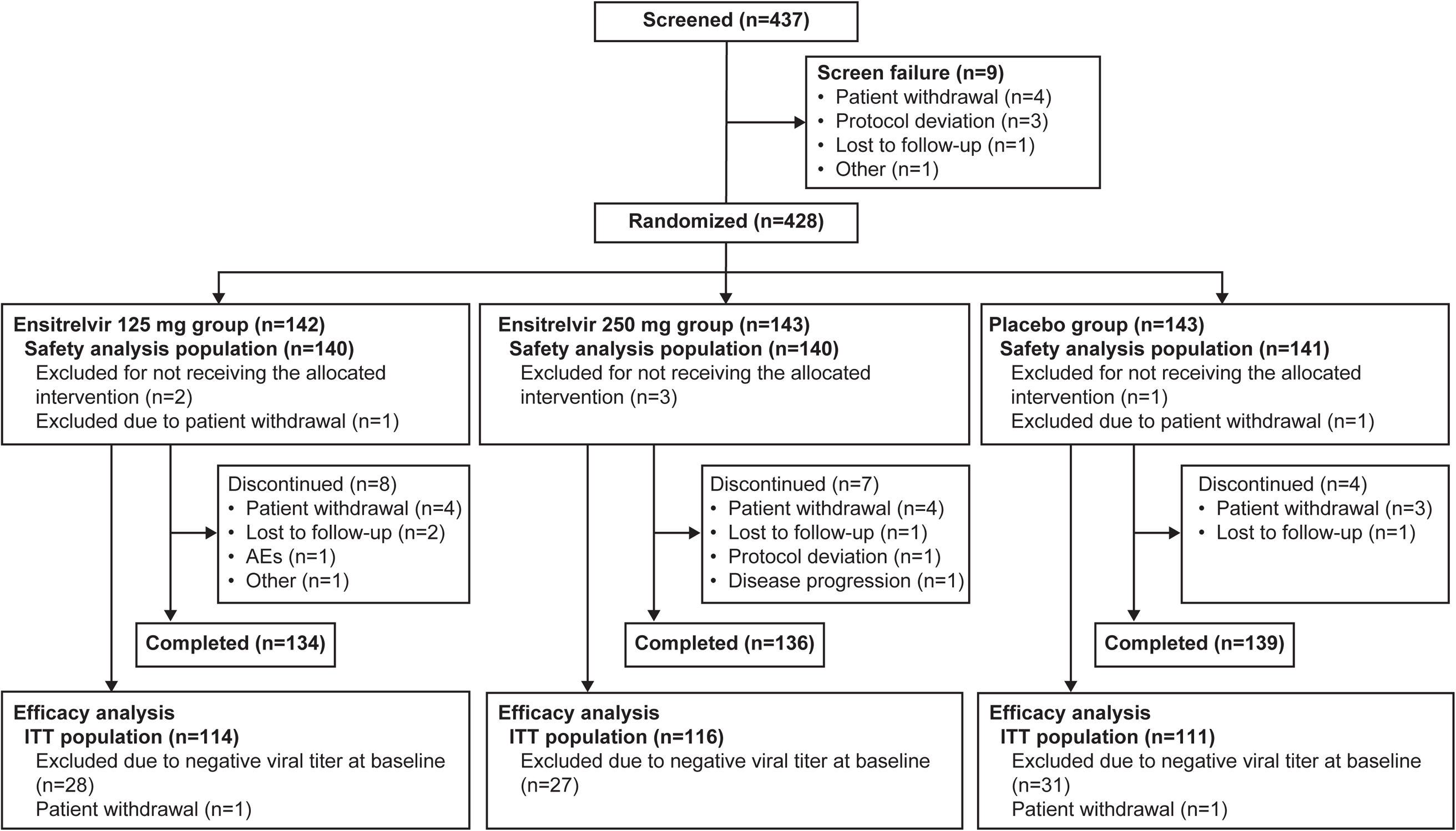
Patient disposition. Some patients were excluded from the analysis populations due to more than one reason. AE, adverse event; ITT, intention-to-treat.

### Demographics and clinical characteristics

The baseline demographics and clinical characteristics of the ITT population are summarized in Table 1. No notable difference was observed in the baseline characteristics across the treatment groups; the mean age was 35.6, 35.3, and 37.3 years, and 1 (0.9%), 2 (1.7%), and 1 (0.9%) patients were aged >65 years in the ensitrelvir 125 mg, ensitrelvir 250 mg, and placebo groups, respectively. A total of 61 (53.5%), 66 (56.9%), and 72 (64.9%) patients in the ensitrelvir 125 mg, ensitrelvir 250 mg, and placebo groups, respectively, were male. Nearly half (ensitrelvir 125 mg, 55 [48.2%]; ensitrelvir 250 mg, 53 [45.7%]; and placebo, 54 [48.6%]) of the patients were randomized within 72 hours of COVID-19 symptom onset, and >80% had received at least 1 dose of mRNA SARS-CoV-2 vaccine (ensitrelvir 125 mg, 97 [85.1%]; ensitrelvir 250 mg, 97 [83.6%]; and placebo, 97 [87.4%]). Among the predefined 12 COVID-19 symptoms (Supplementary Table 2), sore throat (ensitrelvir 125 mg, 65 [57.0%]; ensitrelvir 250 mg, 63 [54.3%]; and placebo, 54 [48.6%]) and cough (ensitrelvir 125 mg, 48 [42.1%]; ensitrelvir 250 mg, 46 [39.7%]; and placebo, 49 [44.1%]) were most frequently observed in all treatment groups.

**Table 1.**
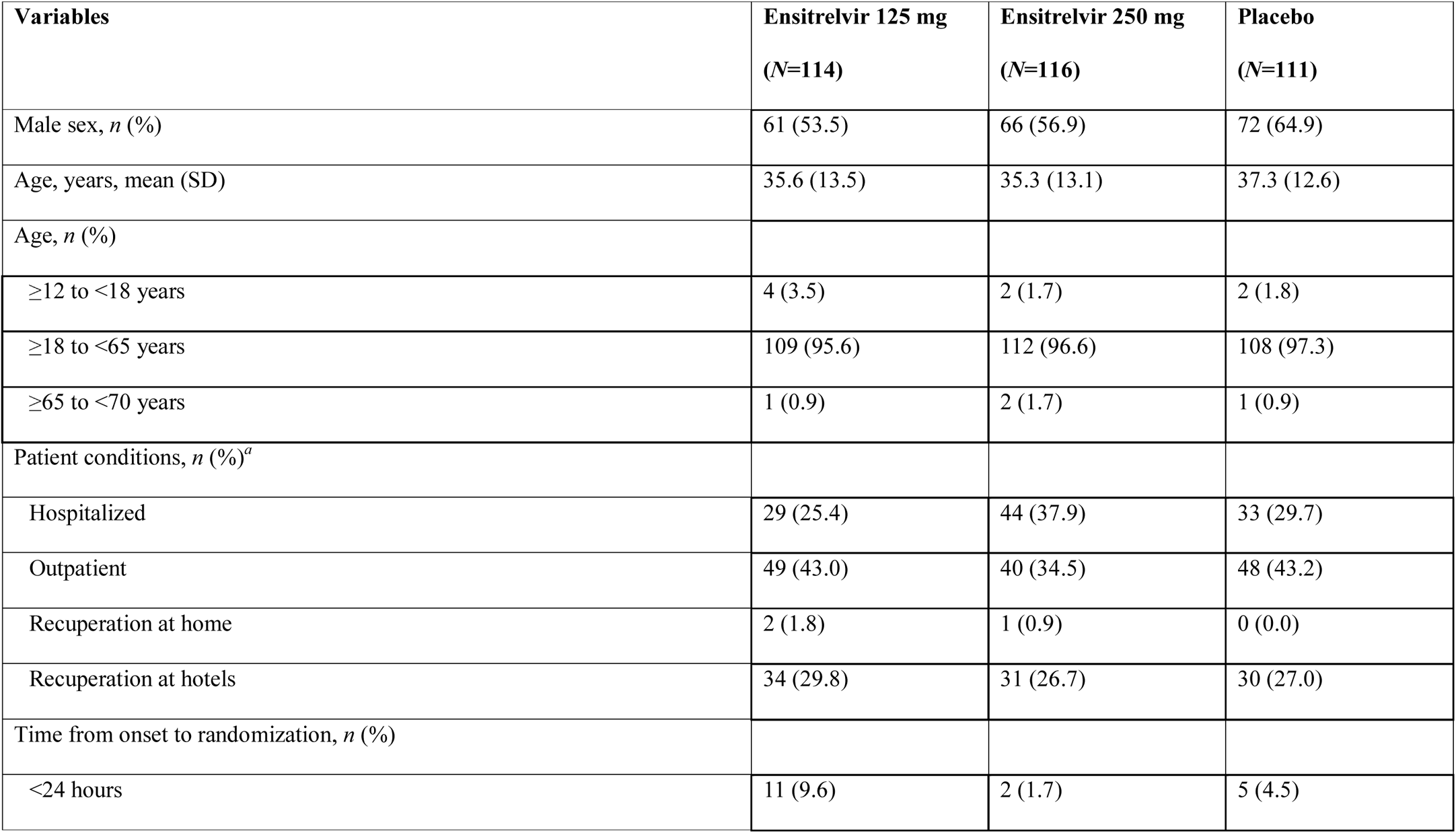
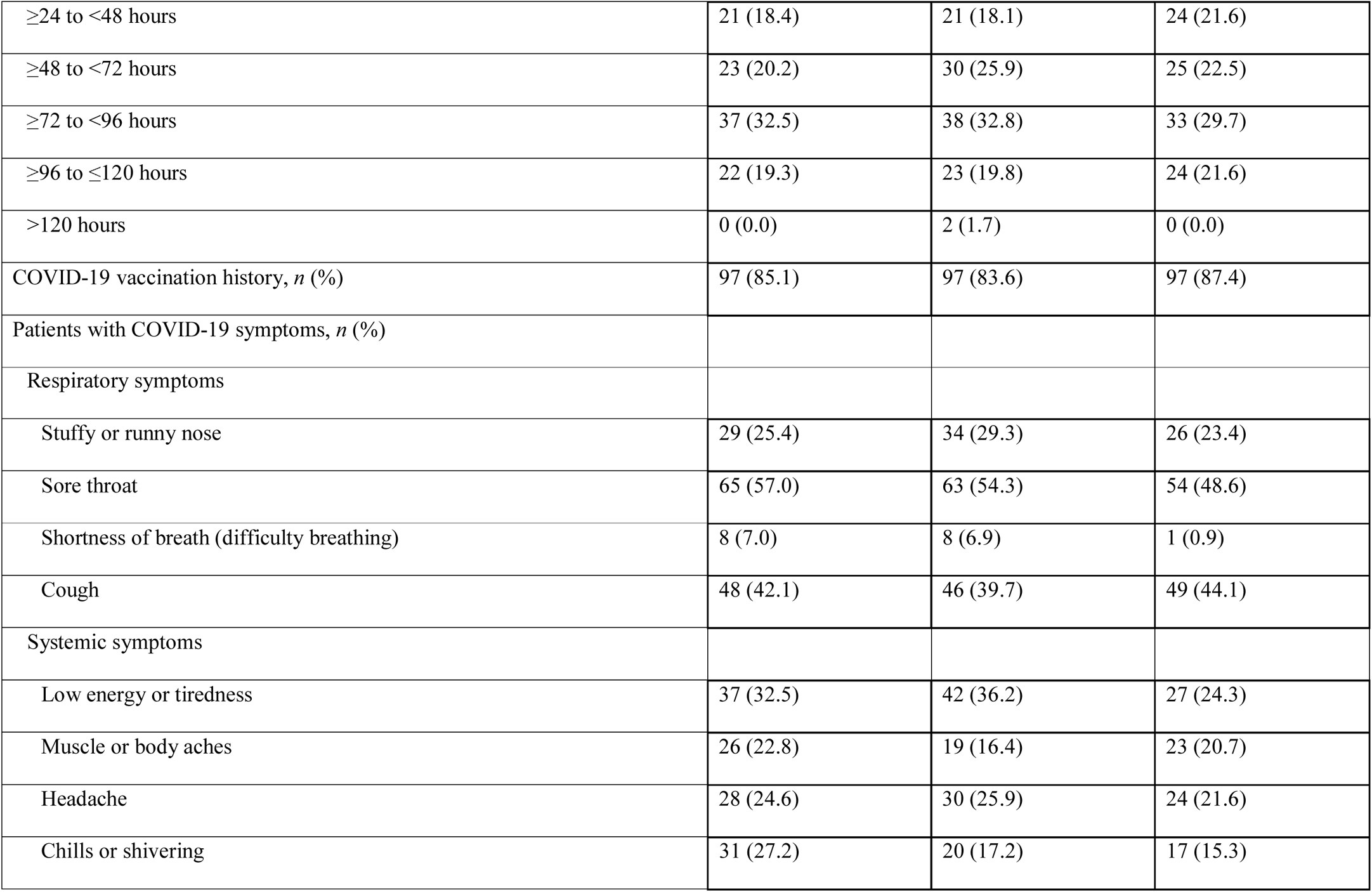
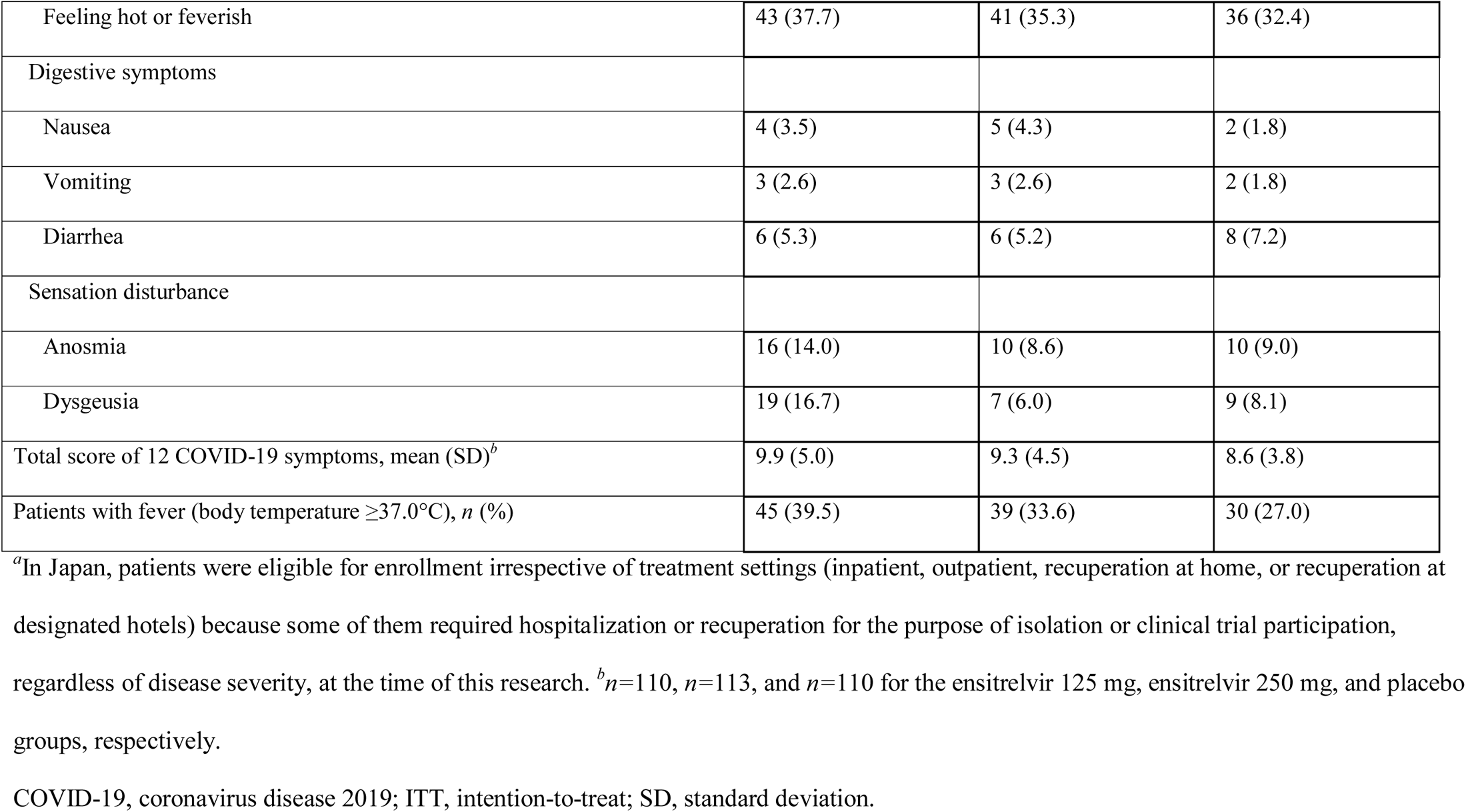
Baseline demographics and clinical characteristics (ITT population)

### Treatment compliance

The treatment compliance rate in the safety population was high across the treatment groups (mean value: ensitrelvir 125 mg, 97.9%; ensitrelvir 250 mg, 99.3%; and placebo, 99.3%).

### Co-primary efficacy endpoint: SARS-CoV-2 viral titer

The mean SARS-CoV-2 viral titer, measured as log_10_ 50% tissue-culture infectious dose ([TCID_50_]/mL), was comparable across groups at baseline (2.5–2.8 log_10_ TCID_50_/mL). In all treatment groups, the SARS-CoV-2 viral titer decreased with time for up to day 4 (after the third drug administration) and remained stable at the levels of lower limit of detection (1.1 log_10_ TCID_50_/mL) until day 21 (Fig. 2a,b). The change from baseline in the SARS-CoV-2 viral titer (log_10_ TCID_50_/mL) on day 4, assessed using analysis of covariance (ANCOVA), was significantly greater with ensitrelvir 125 mg and 250 mg (least-square [LS] mean [standard error (SE)], −1.49 [0.04]; difference from placebo, −0.41 [95% confidence interval (CI), −0.51 to −0.31]; *P*<0.0001) versus placebo (LS mean [SE], −1.08 [0.04]). The change from baseline in the SARS-CoV-2 viral titer on day 4 was significantly greater with ensitrelvir 125 mg and 250 mg versus placebo irrespective of COVID-19 vaccination history or time from COVID-19 onset to randomization (Supplementary Table 3).

**Fig. 2:**
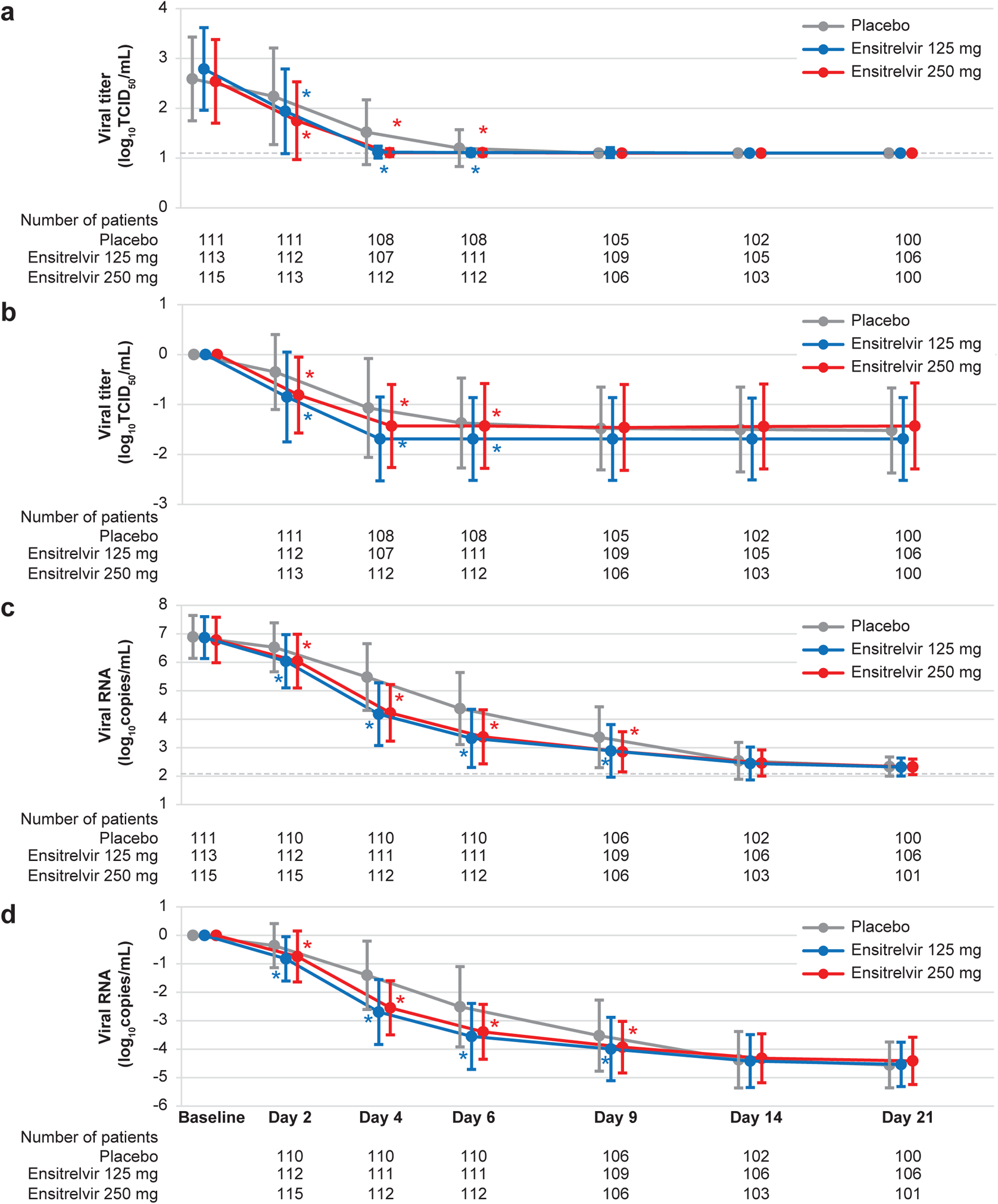
a, Absolute values and b, change from baseline in SARS-CoV-2 viral titer; c, absolute values and d, change from baseline in SARS-CoV-2 RNA level (ITT population). Data are presented as mean±SD. \**P*<0.05 versus placebo. Dotted lines indicate the lower limit of detection for SARS-CoV-2 viral titer (1.1 log_10_ TCID_50_/mL) and lower limit of quantification for SARS-CoV-2 RNA level (2.08 log_10_ copies/mL). ITT, intention-to-treat; SARS-CoV-2, severe acute respiratory syndrome coronavirus 2; SD, standard deviation; TCID_50_, 50% tissue-culture infectious dose.

### Co-primary efficacy endpoint: Total scores for 12 COVID-19 symptoms

The mean total score of the predefined 12 COVID-19 symptoms showed a decreasing trend with time after treatment initiation in all groups (Extended Data Fig. 1). There was no significant difference in the time-weighted average change from baseline up to 120 hours in the total score of the 12 COVID-19 symptoms between the ensitrelvir 125 mg or 250 mg group and the placebo group (Table 2).

**Table 2.** Time-weighted average change from baseline up to 120 hours in the total score of 12 COVID-19 symptoms (ITT population)

| Statistics | Ensitrelvir 125 mg<br>(N=114) | Ensitrelvir 250 mg<br>(N=116) | Placebo<br>(N=111) |
| --- | --- | --- | --- |
| N | 109 | 113 | 110 |
| Mean (SD) change from baseline | -5.95 (4.02) | -5.42 (3.70) | -4.92 (3.25) |
| LS mean (SE) change from baseline assessed by ANCOVA | -5.37 (0.24) | -5.17 (0.23) | -5.12 (0.24) |
| LS mean (SE) difference in change from baseline versus placebo | -0.24 (0.30) | -0.04 (0.29) | – |
| 95% CI | -0.83 to 0.34 | -0.62 to 0.53 | – |
| P-value | 0.4171 | 0.8806 | – |

ANCOVA, analysis of covariance; CI, confidence interval; COVID-19, coronavirus disease 2019; ITT, intention-to-treat; LS, least squares; SD, standard deviation; SE, standard error.

### SARS-CoV-2 viral RNA level

The SARS-CoV-2 RNA level decreased with time in all groups (Fig. 2c,d). The change from baseline in SARS-CoV-2 RNA level (log_10_ copies/mL) on day 4, assessed using ANCOVA, was significantly greater with ensitrelvir 125 mg (LS mean [SE], −2.58 [0.11]; difference from placebo, −1.30 [95% CI, −1.57 to −1.03]; *P*<0.0001) and 250 mg (LS mean [SE], −2.49 [0.11]; difference from placebo, −1.21 [95% CI, −1.48 to −0.94]; *P*<0.0001) versus placebo (LS mean [SE], −1.28 [0.11]). Similarly, the change from baseline in SARS-CoV-2 RNA level was significantly greater in the ensitrelvir groups versus the placebo group on days 2, 6, and 9.

### Time to first negative SARS-CoV-2 viral titer

The time to first negative SARS-CoV-2 viral titer (infectious viral clearance) was significantly shorter with ensitrelvir 125 mg (51.3 hours; difference of median time from placebo, −40.6 hours; *P*<0.0001) and 250 mg (62.1 hours; difference of median time from placebo, −29.8 hours; *P*<0.0001) versus placebo (91.9 hours) (Fig. 3).

**Fig. 3:**
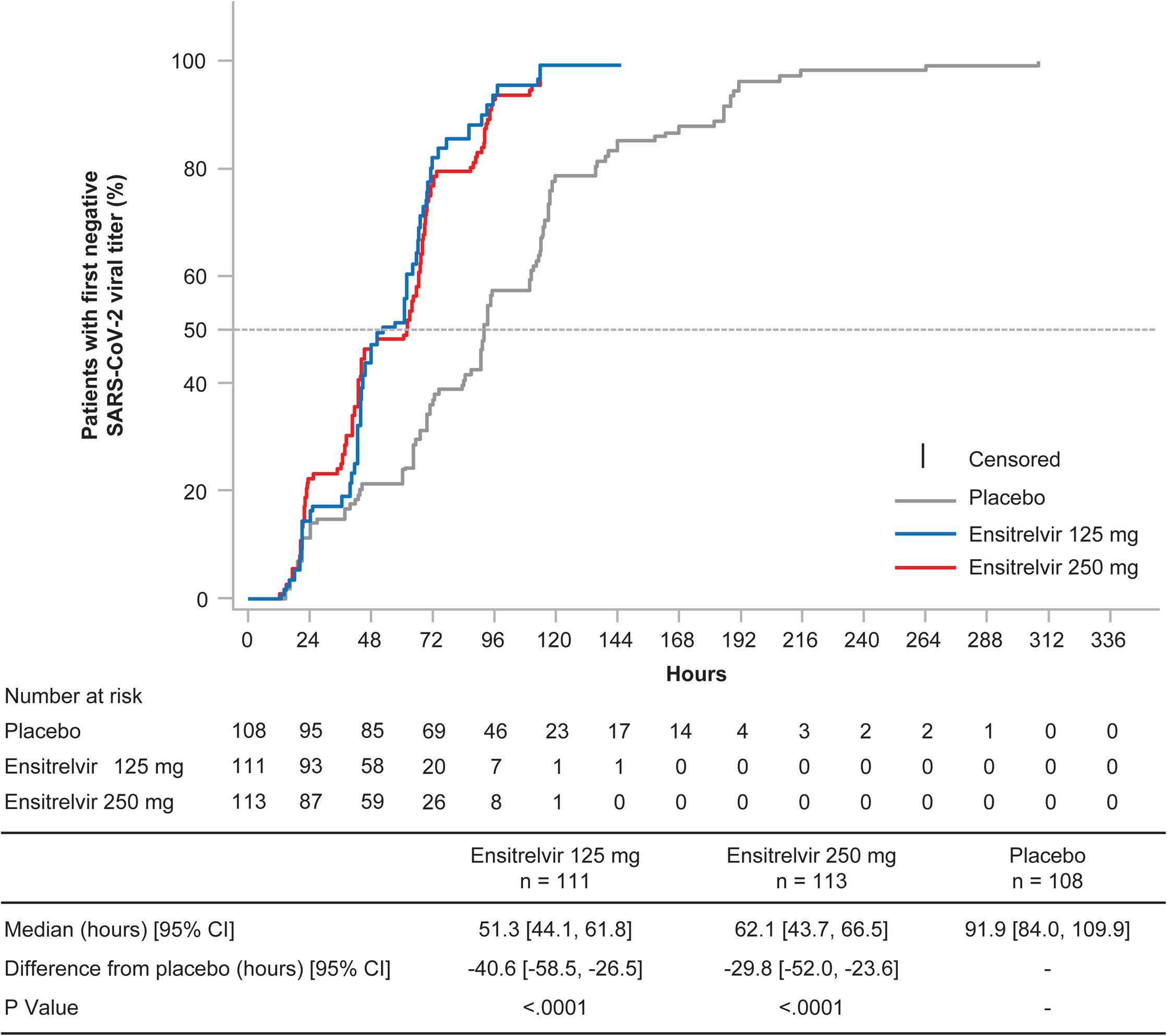
Time to first negative SARS-CoV-2 viral titer (viral clearance; ITT population). CI, confidence interval; ITT, intention-to-treat; SARS-CoV-2, severe acute respiratory syndrome coronavirus 2.

### Proportion of patients with a positive SARS-CoV-2 viral titer

The proportion of patients with a positive SARS-CoV-2 viral titer decreased with time in all treatment groups; on day 4, it was significantly lower with ensitrelvir 125 mg (1.9%, *P*<0.0001) and 250 mg (5.4%, *P*<0.0001) versus placebo (50.0%), with a reduction of approximately 90%. Similarly, the proportion of patients with a positive SARS-CoV-2 viral titer was significantly lower with ensitrelvir 125 mg and 250 mg versus placebo on day 6 (Extended Data Fig. 2).

### Subtotal scores for the 12 COVID-19 symptoms

The mean changes from baseline in the subtotal scores of the 12 COVID-19 symptoms are depicted in Extended Data Fig. 3. In addition to the preplanned subtotal scores (acute symptoms, main clinical symptoms, respiratory symptoms, systemic symptoms, and digestive symptoms), a composite subtotal score of respiratory symptoms and feverishness was evaluated as a post hoc analysis. The time-weighted average change from baseline up to 120 hours was significantly greater with ensitrelvir versus placebo in the subtotal scores for acute symptoms (250 mg group, *P*=0.0070), main clinical symptoms (250 mg group, *P*=0.0149), respiratory symptoms (125 mg and 250 mg groups, *P*=0.0153 and 0.0033, respectively), and the composite of respiratory symptoms and feverishness (125 mg and 250 mg groups, *P*=0.0164 and 0.0039, respectively). In contrast, no significant difference was observed between the ensitrelvir and placebo groups in the time-weighted average change for systemic symptoms and digestive symptoms (Supplementary Table 4). The anosmia and dysgeusia scores generally showed a transient increase from baseline, suggesting a delayed onset of these symptoms in patients with COVID-19. The change from baseline in the anosmia and dysgeusia scores was lower with ensitrelvir 125 mg and 250 mg versus placebo (Extended Data Fig. 4).

No significant difference was observed in the time to first improvement of COVID-19 symptoms (median [95% CI] hours: 28.0 [21.5 to 36.6], 27.8 [24.6 to 40.0], and 36.6 [28.0 to 40.8] for ensitrelvir 125 mg, ensitrelvir 250 mg, and placebo, respectively).

### COVID-19 exacerbation

After treatment initiation, 1 vaccinated patient in the ensitrelvir 250 mg group was rated as ≥3 (corresponding to hospitalization or death) by the investigator on an 8-point ordinal scale for disease exacerbation. None of the patients in the ensitrelvir 125 mg and placebo groups recorded ≥3 on the disease exacerbation scale.

### Safety

Overall, 48 (34.3%), 60 (42.9%), and 44 (31.2%) patients in the ensitrelvir 125 mg, ensitrelvir 250 mg, and placebo groups, respectively, reported treatment-emergent adverse events (TEAEs), most of which were mild in severity. Treatment-related adverse events (AEs) were observed in 19 (13.6%), 31 (22.1%), and 7 (5.0%) patients in the ensitrelvir 125 mg, ensitrelvir 250 mg, and placebo groups, respectively, a majority of which were resolved without sequelae. A decrease in high-density lipoprotein (HDL) levels was most frequently reported as a TEAE across groups (ensitrelvir 125 mg, 31 [22.1%] patients; ensitrelvir 250 mg, 40 [28.6%] patients; and placebo, 5 [3.5%] patients) and was the most common treatment-related AE in both ensitrelvir groups (125 mg, 13 [9.3%] patients; 250 mg, 22 [15.7%] patients). No TEAEs leading to death were reported during the study (Table 3).

**Table 3.**
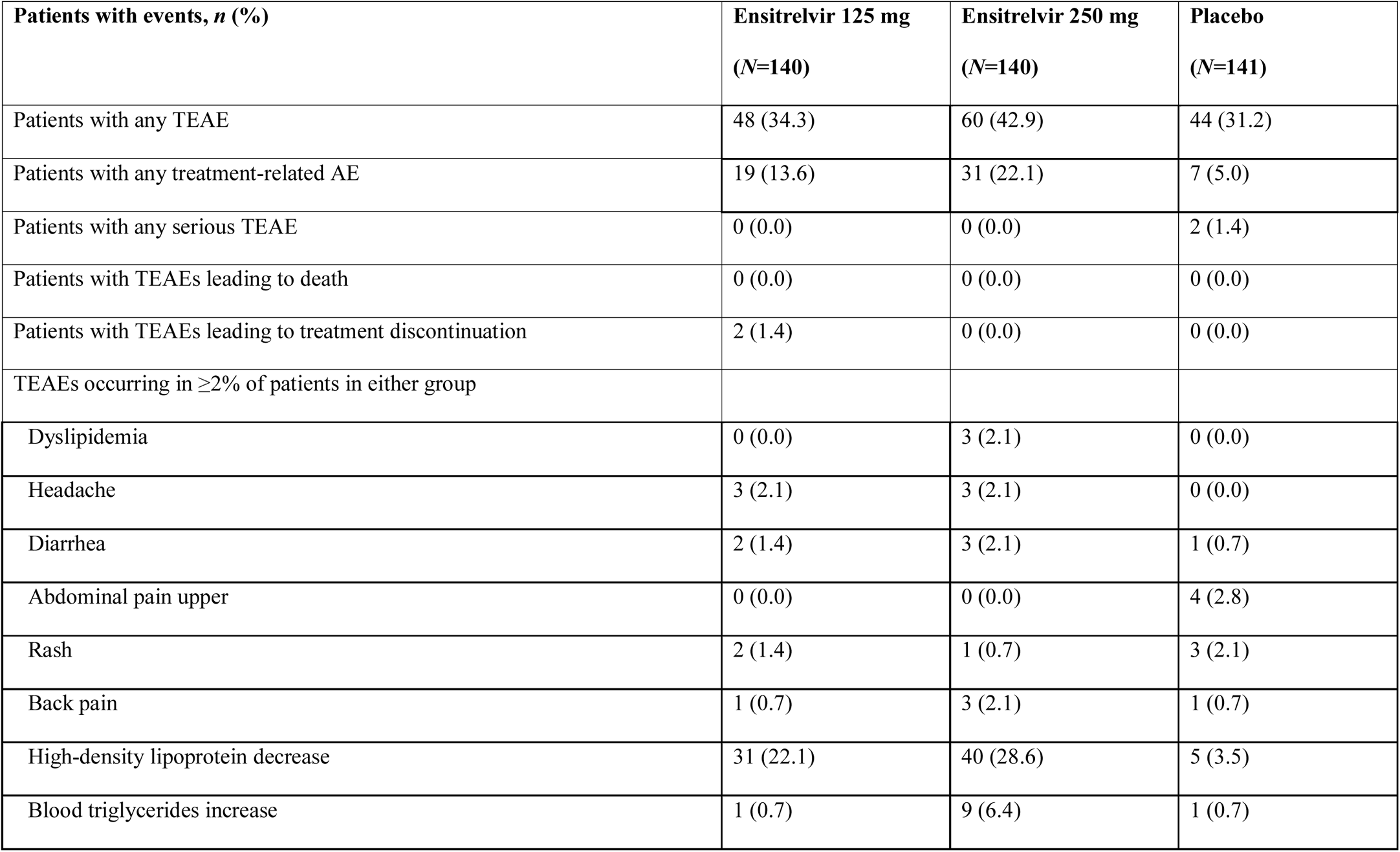
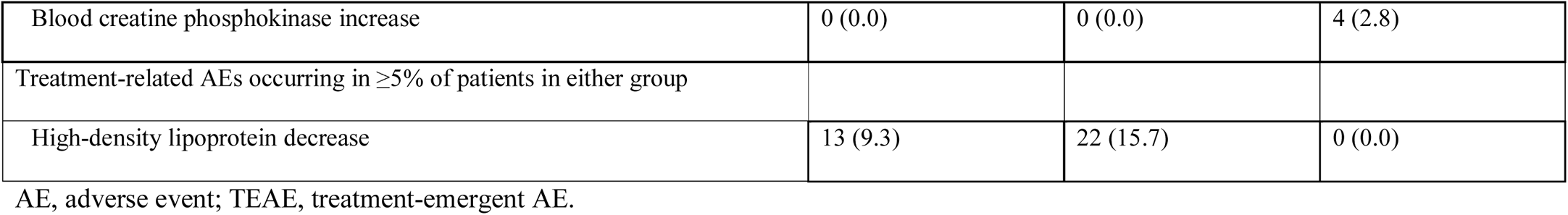
Summary of TEAEs (safety analysis population)

Two patients in the placebo group had serious TEAEs (thoracic vertebral fracture [recovered without sequelae] and facial paralysis [recovering]), both of which were determined as not treatment related. Another 2 patients in the ensitrelvir 125 mg group discontinued the treatment due to TEAEs. All 3 TEAEs leading to treatment discontinuation (mild eczema on day 2 in 1 patient and moderate nausea and mild headache on day 2 in 1 patient) were judged as treatment related by the investigator, and both patients recovered without sequelae after drug discontinuation.

A dose-dependent, transient change in the HDL cholesterol, triglyceride, total bilirubin, and iron levels was observed on day 6 in the ensitrelvir groups (Extended Data Fig. 5). These changes were asymptomatic and resolved without additional treatment.

No notable difference was observed with ensitrelvir treatment versus placebo in the haptoglobin, reticulocyte, or low-density lipoprotein levels, and no laboratory or clinical signs of hemolysis were observed. None of the patients showed serum bilirubin increase occurring concurrently with liver enzyme elevation.

## Discussion

This phase 2b part of a randomized, double-blind, placebo-controlled, phase 2/3 trial of ensitrelvir in patients with mild-to-moderate COVID-19 was conducted in early 2022 during the SARS-CoV-2 Omicron epidemic. Based on the results of the phase 2a part conducted during the SARS-CoV-2 Delta epidemic^20^, which showed a statistically significant reduction in the SARS-CoV-2 viral titer and an improving trend in the total score of the 12 COVID-19 symptoms after ensitrelvir treatment, we used these outcomes as the co-primary endpoints. On day 4, ensitrelvir treatment significantly reduced the SARS-CoV-2 viral titer versus placebo. There was no significant difference from placebo in the time-weighted average change from baseline up to 120 hours in the total score of the 12 COVID-19 symptoms. Of note, because of the difference in the predominant SARS-CoV-2 variants, virologic characteristics and clinical disease course observed in this phase 2b part may not be consistent with those observed in the phase 2a part or previous anti–COVID-19 clinical trials and may affect the interpretation of the virologic and clinical efficacy of ensitrelvir.

In the current phase 2b part, virologic efficacy was assessed using the change in SARS-CoV-2 viral titer from baseline to day 4 (co-primary endpoint) and SARS-CoV-2 RNA (endpoint commonly used in clinical studies for anti–COVID-19 treatments^14, 16, 21^). The rapid and significant reduction in the SARS-CoV-2 viral titer and viral RNA following ensitrelvir treatment is consistent with the findings of the previous phase 2a part^20^, but the level of viral titer reduction from baseline to day 4 was lower in the current phase 2b part (−1.49 log_10_ TCID_50_/mL) than in the phase 2a part (−2.81 log_10_ TCID_50_/mL with ensitrelvir 250 mg)^20^. These differences in findings may be attributed to the lower baseline titer recorded in the phase 2b part than in the phase 2a part (2.5–2.8 versus 3.3–3.7 log_10_ TCID_50_/mL), as well as other unidentified differences in viral characteristics. Indeed, previous research findings indicate different characteristics between the Omicron and Delta variants in the infectivity or peak viral titer measured in Vero-E6 cells^22^ and peak viral load and viral clearance^23^, despite the consistent antiviral efficacy of ensitrelvir against the Omicron variant shown in preclinical research^17–19^. The difference from placebo in the reduction in SARS-CoV-2 RNA observed from baseline to day 4 in the current phase 2b part (−1.30 and −1.21 log_10_ copies/mL for ensitrelvir 125 and 250 mg, respectively) appeared greater than that observed with molnupiravir 800 mg (day 5, −0.547 log_10_ copies/mL)^21^, nirmatrelvir 300 mg in combination with ritonavir 100 mg (day 5, −0.868 log_10_ copies/mL)^14^, or casirivimab/imdevimab 2400 mg (day 7, −0.86 log_10_ copies/mL)^16^. The majority of patients enrolled in the current phase 2b part were vaccinated, whereas previous anti–COVID-19 treatments were evaluated in unvaccinated patients at risk of severe disease^13, 14^. Although no direct comparison with previous clinical trials is feasible due to differences in study settings, our results support the promising efficacy of ensitrelvir in real-world clinical settings. Moreover, no notable viral rebound (increase in viral titer or RNA) was observed on day 14 or 21 of ensitrelvir treatment in contrast to the findings observed with nirmatrelvir treatment (https://www.fda.gov/media/155194/download). The absence of viral rebound may be attributed to a longer half-life of ensitrelvir (51.4 hours; R. Shimizu et al., unpublished data) versus nirmatrelvir 250 mg in combination with ritonavir 100 mg (approximately 6.5 hours)^24^, which should be further assessed in large-scale, confirmatory trials.

Clinical efficacy, such as a symptom relief, is a pivotal part of COVID-19 clinical trials because patients present with diverse symptoms^25^ and clinical symptoms may differ depending on the epidemic variants. Although the decreasing trend in the total 12 COVID-19 symptom scores is consistent with the findings from the phase 2a part^20^, the results of the current phase 2b part indicate differences in the symptom characteristics between the Omicron and Delta variant infections. Among the 12 symptoms assessed, respiratory symptoms and feverishness were recorded in many patients enrolled in the phase 2b part, whereas systemic and digestive symptoms were relatively less observed at baseline. These findings can be explained by an epidemiology report in Japan, where the most commonly reported COVID-19 symptoms in patients infected with the Omicron variant were respiratory symptoms, such as cough (46.0%), sore throat (33.8%), runny nose (18.0%), and fever (30.9%) (https://www.niid.go.jp/niid/ja/2019-ncov/2484-idsc/10969-covid19-72.html; website in Japanese). The absence of significant intergroup difference in the total score of the 12 COVID-19 symptoms can be attributed to the low baseline subtotal scores recorded for some patients (e.g., digestive symptoms) in the current phase 2b part. Interestingly, ensitrelvir treatment improved respiratory symptoms or a composite outcome of respiratory symptoms and feverishness versus placebo, which suggests the potential clinical benefits of this antiviral in patients infected with the Omicron variant.

The current phase 2b part of the study found no clear dose response, which is consistent with the findings of the phase 2a part^20^. Pharmacokinetic data for 5-day ensitrelvir 125 mg treatment (375 mg on day 1) derived from the phase 1 study of ensitrelvir (maximum plasma concentration, 30.4 µg/mL; area under the curve, 598.3 µg·hr/mL) suggest that this regimen is sufficient to achieve a reduction in SARS-CoV-2 in humans (R. Shimizu et al., unpublished data). Ensitrelvir 125 mg (375 mg on day 1) is thus considered the optimal dose for achieving an antiviral effect while minimizing drug exposure.

Infection with the Omicron variant causes less severe disease than infection with the Delta variant^26–28^, which may be attributed to altered infectivity and attenuated lung disease with the Omicron variant observed in rodent models^29^ or immunity acquired via previous viral exposure through vaccination and/or past infections^27, 28^. In the current phase 2b part, >95% of patients receiving placebo showed symptom improvement by day 5, and the median time to first improvement of the 12 COVID-19 symptoms (27.8–36.6 hours) was shorter than that observed in the previous phase 2a part (36.7–55.2 hours), which may reflect less disease severity of infection with the Omicron variant. New clinical endpoints that can assess the clinical symptoms of COVID-19 are warranted.

In addition to treatment options for patients at risk of severe disease, effective antivirals that can rapidly reduce SARS-CoV-2 are warranted because patients with asymptomatic or mild-to-moderate COVID-19 may shed infectious viruses for up to 10 days and may be a source for viral transmission^30^. Early treatment initiation for mild-to-moderate COVID-19 is necessary in view of the disease characteristics and to gain maximum benefits from pharmacological therapies (https://www.kansensho.or.jp/uploads/files/topics/2019ncov/covid19_drug_220218.pdf; website in Japanese). Moreover, long-term complications after COVID-19, such as loss of taste and/or smell, fatigue, headache, attention disorder, hair loss, and dyspnea, can occur in the relatively healthy population (e.g., young adults or patients with mild disease) and contribute toward persistent and significant health issues^31, 32^. Although the exact mechanism of the onset of long-term COVID-19 complications is unknown, rapid SARS-CoV-2 reduction with ensitrelvir treatment may be effective in suppressing these complications, which should be assessed in an optional, exploratory period (days 28 to 337) of the current phase 2/3 study.

Similar to the previous phase 1 study (R. Shimizu et al., unpublished data) and phase 2a part of the current study^20^, a transient change in the HDL cholesterol and triglyceride levels was observed in patients treated with ensitrelvir. In addition, total bilirubin and iron levels transiently increased with ensitrelvir treatment. The underlying causes of these laboratory findings and their potential relationship with ensitrelvir treatment warrant further investigation.

This study has some limitations. The number of elderly patients (aged ≥65 years) was limited; thus, data analysis for this population was not feasible. As the risk of severe COVID-19 increases with age^33^, the safety and efficacy of ensitrelvir in a wide range of patients with COVID-19 should be further assessed in the phase 3, multinational, randomized, placebo-controlled study, SCORPIO-HR (ClinicalTrials.gov identifier: NCT05305547). Moreover, as new variants may arise in the future, continuous monitoring and assessment of ensitrelvir activity against new VOCs would be required. In addition to the preclinical antiviral activity of ensitrelvir against a variety of SARS-CoV-2 variants^17^, evidence derived in clinical trial settings is awaited.

In conclusion, 5-day, once daily, oral ensitrelvir treatment demonstrated rapid and favorable antiviral efficacy with an acceptable safety profile in patients with mild-to-moderate COVID-19, a majority of whom had been vaccinated. Ensitrelvir treatment improved respiratory symptoms and feverishness, which were common COVID-19 symptoms during the Omicron variant epidemic. The results support further clinical development of ensitrelvir for mild-to-moderate COVID-19.

## Methods

### Study design

Patients with mild-to-moderate COVID-19 were randomized (1:1:1) to orally receive ensitrelvir fumaric acid (375 mg on day 1, followed by 125 mg on days 2 through 5, or 750 mg on day 1, followed by 250 mg on days 2 through 5) or matching placebo once daily without dose modification and followed up until day 28.

This study was conducted in accordance with the principles of the Declaration of Helsinki, Good Clinical Practice guidelines, and other applicable laws and regulations. The study was reviewed and approved by the institutional review boards of all participating institutions listed in Supplementary Table 1. All patients or their legally acceptable representatives provided written informed consent.

### Patients

Patients aged 12 to 69 years who tested positive for SARS-CoV-2 (assessed by SARS-CoV-2 antigen or nucleic acid detection testing) within 120 hours prior to randomization were eligible for study enrollment. To avoid excess drug exposure, patients aged <20 years should have recorded a body weight of ≥40 kg at enrollment. Patients should have at least one moderate or severe symptom or worsening of an existing symptom among the 12 COVID-19 symptoms (stuffy or runny nose, sore throat, shortness of breath, cough, low energy or tiredness, muscle or body aches, headache, chills or shivering, feeling hot or feverish, nausea, vomiting, or diarrhea; Supplementary Table 2) based on the US Food and Drug Administration (US FDA) guidance (https://www.fda.gov/regulatory-information/search-fda-guidance-documents/assessing-covid-19-related-symptoms-outpatient-adult-and-adolescent-subjects-clinical-trials-drugs) and a symptom duration of ≤120 hours. In Japan, patients were eligible for enrollment irrespective of treatment settings (inpatient, outpatient, recuperation at home, or recuperation at designated hotels) because some of them required hospitalization or recuperation for the purpose of isolation or clinical trial participation, regardless of disease severity, at the time of this research.

The key exclusion criteria included the following: an awake oxygen saturation of ≤93% (room air); oxygen administration; likely worsening of COVID-19 within 48 hours from randomization in the opinion of the investigator; suspected active and systemic infections requiring treatment (except for COVID-19); current or chronic history of moderate or severe liver disease, known hepatic or biliary abnormalities (except for Gilbert’s syndrome or asymptomatic gallstones), or moderate-to-severe kidney disease; pregnant, possibly pregnant, or breastfeeding women; and blood donation (≥400 mL within 12 weeks or ≥200 mL within 4 weeks prior to enrollment). Patients who had used drugs for SARS-CoV-2 infection within 7 days prior to randomization or a strong cytochrome P450, family 3, subfamily A (CYP3A) inhibitor or inducer or St. John’s wort products within 14 days prior to randomization were also excluded.

### Randomization and blinding

Randomization of patients was performed through an interactive response technology system using time from the onset of COVID-19 to randomization (<72 hours/≥72 hours) and the first COVID-19 vaccination (Yes/No) as stratification factors. All patients and study staff were blinded to the treatment, except for designated persons at the sponsor and contract research organization in charge of statistical analyses. When all patients completed the day 6 assessment, the study sponsor was unblinded to intervention allocation to allow for an early-stage evaluation of the efficacy and safety data. Investigators, patients, and other study staff remained blinded until the completion of the 28-day follow-up period. Emergency unblinding per the investigator’s request was allowed only in the event of AEs to determine appropriate therapy for the patient.

### Treatment

Patients received allocated study drugs orally (ensitrelvir fumaric acid 125 mg, 250 mg, or placebo tablets), which were indistinguishable in appearance, labeling, and packaging. Prohibited concomitant medications (study initiation to day 28 or study discontinuation) included drugs for the treatment of SARS-CoV-2 infection; antiviral, antibacterial, or antifungal drugs (except for topical use); antipyretic analgesics other than acetaminophen; antitussives and expectorants; combination cold remedy; and CYP3A substrates. Drugs prohibited for use from study initiation to 10 days after the last study drug administration (or study discontinuation) included strong CYP3A inhibitors or inducers, strong P-glycoprotein or breast cancer resistance protein inhibitors, and other transporter substrates. Treatment was discontinued when liver function abnormalities, pregnancy, COVID-19 exacerbation, or serious or intolerable AEs were observed.

### Outcomes and assessments

The primary virologic outcome was change from baseline (day 1, before drug administration) in the SARS-CoV-2 viral titer on day 4 of treatment. The primary clinical outcome was time-weighted average change from baseline up to 120 hours in the total score of COVID-19 symptoms. The co-primary endpoint was constructed by the primary virologic and clinical outcomes. The secondary outcomes included the SARS-CoV-2 viral titer and viral RNA level (absolute values and change from baseline) up to day 21, time to first negative SARS-CoV-2 viral titer (infectious viral clearance), proportion of patients with positive viral titers, subtotal scores of the 12 COVID-19 symptoms (acute symptoms, main clinical symptoms, respiratory symptoms, systemic symptoms, and digestive symptoms; Supplementary Table 2), time to first improvement of COVID-19 symptoms (Supplementary Table 2), and COVID-19 exacerbations assessed by the investigator. Additionally, post hoc analyses were performed to assess a composite subtotal score (respiratory symptoms and feverishness) for the 12 COVID-19 symptoms.

Nasopharyngeal swabs were collected from the patients on days 1 (before drug administration), 2 to 6 (days 3 and 5 as optional), 9, 14, and 21 (or study discontinuation), and the SARS-CoV-2 viral titers and RNA levels were centrally measured at Shionogi TechnoAdvance Research (Osaka, Japan) and Viroclinics (Rotterdam, Netherlands), respectively. For the assessment of COVID-19 symptoms, patients rated each symptom on a 4-point or 3-point scale (Supplementary Table 2) and recorded the scores in a diary twice daily (morning and evening) until day 9 and once daily (evening) from days 10 to 21. Additionally, patients’ COVID-19 exacerbation was assessed by the investigator using an 8-point ordinal scale (0=Asymptomatic; 1=Symptomatic, no limitation of activities; 2=Symptomatic, limitation of activities; 3=Hospitalized, no oxygen therapy; 4=Hospitalized, with oxygen therapy [<5 L/min]; 5=Hospitalized, with oxygen therapy [≥5 L/min]; 6=Hospitalized, with ventilation; 7=Death) on day 1 (before drug administration) and days 2, 4, 6, 9, 14, 21, and 28 (or study discontinuation).

Safety was assessed by the occurrence of TEAEs, that is, any AEs reported after the initiation of study intervention. All AEs were coded and classified using Medical Dictionary for Regulatory Activities version 24.0. Laboratory tests, vital sign measurements, and electrocardiography were additionally performed throughout the study period. All safety data were evaluated by an independent data and safety monitoring board. Pregnancy tests were performed for women with childbearing potential on days 1 (before drug administration) and 28 (or study discontinuation). Additional pregnancy tests at the investigator’s discretion were permitted.

### Statistical analyses

Based on the interim evaluation of the phase 2a part of this study (Shionogi & Co., Ltd., unpublished data), the difference in time-weighted average change in the total score of COVID-19 symptoms from baseline up to 120 hours between the ensitrelvir and placebo groups was assumed to be −1, with a standard deviation (SD) of 2.6. A total of 108 patients per group (324 in total) were required to detect this difference with 80% power using a two-sample t-test at a one-sided significance level of 0.025. The same sample size was required to detect the intergroup difference in change from baseline in the SARS-CoV-2 viral titer on day 4 with 99.9% power using a two-sample t-test at a one-sided significance level of 0.025, assuming a difference between the ensitrelvir and placebo groups of −0.5 log_10_ TCID_50_/mL and an SD of 0.7 log_10_ TCID_50_/mL. Considering a dropout rate of 25% (proportion of patients with a negative SARS-CoV-2 viral titer at baseline), the final sample size was set at 145 patients per group (435 in total).

All randomized patients with a positive SARS-CoV-2 viral titer (≥1.1 log_10_ TCID_50_/mL) at baseline were included in the ITT population. All randomized patients who received at least one dose of the study drug were included in the safety analysis population.

A prespecified statistical hypothesis was set, wherein both virologic and clinical efficacy are confirmed if differences in the co-primary endpoints of virologic and clinical outcomes between the ensitrelvir (125 mg or 250 mg) and placebo groups are statistically significant at a one-sided significance level of 0.025. To control the type I error for multiplicity of testing that occurred in pairwise comparisons between each of the ensitrelvir dose groups and the placebo group, a fixed sequence procedure was applied to the primary virologic and clinical outcomes, where the statistical tests were performed in the order of ensitrelvir 125 mg versus placebo and ensitrelvir 250 mg versus placebo. Other statistical tests were performed at a two-sided significance level of 0.05.

An ANCOVA model was constructed for the primary analysis of the primary endpoints for pairwise comparison of each efficacy outcome between each of the ensitrelvir groups and the placebo group using baseline values, time from COVID-19 onset to randomization (<72 or ≥72 hours), and COVID-19 vaccination history (Yes or No) as covariates. LS mean, difference from placebo, and 95% CI were calculated based on the ANCOVA model. Time-weighted average change from baseline up to 120 hours in the total score of the 12 COVID-19 symptoms was calculated by dividing the area under the curve up to 120 hours for change from baseline in the total score of the 12 COVID-19 symptoms by hours from the start of first drug administration. A Kaplan-Meier curve was plotted for the analyses of time to first negative SARS-CoV-2 viral titer and time to first improvement of COVID-19 symptoms, and the median time was compared between each of the ensitrelvir groups and the placebo group using a log-rank test stratified by time from COVID-19 onset to randomization and COVID-19 vaccination history. The proportion of patients with a positive SARS-CoV-2 viral titer was compared between each of the ensitrelvir groups and the placebo group using the Mantel-Haenszel test, stratified by time from COVID-19 onset to randomization and SARS-CoV-2 vaccination history.

No imputation was performed for missing data. All analyses were performed using SAS version 9.4 (SAS Institute, Inc., Cary, NC, USA).

## Data availability

Shionogi & Co., Ltd. is committed to disclosing the synopses and results of its clinical trials and sharing the clinical trial data with researchers on reasonable request. For further details, please refer to the websites of Shionogi & Co., Ltd. (https://www.shionogi.com/shionogi/global/en/company/policies/shionogi-group-clinical-trial-data-transparency-policy.html) and Vivli (https://vivli.org/).

## Supporting information

Supplementary Tales 1 to 4

## Acknowledgments

The authors and research team thank all the patients involved in this study and Masahiro Kinoshita and Satoshi Kojima (Shionogi & Co., Ltd.) for preparing technical support documents and an earlier version of the manuscript draft. Support for study monitoring and data management was provided by EPS Corporation and funded by Shionogi & Co., Ltd. Medical writing and editorial assistance was provided by Mami Hirano, MS, of Cactus Life Sciences (part of Cactus Communications) and funded by Shionogi & Co., Ltd. All authors retained full ownership of the manuscript content and approved the final draft for submission.

This study was sponsored by Shionogi & Co., Ltd., and financially supported by the Organization of the Ministry of Health, Labour and Welfare. Employees of Shionogi & Co., Ltd., participated in and approved the design and conduct of the study; wrote the protocol; and were involved in the collection, management, analysis, and interpretation of data. Institutional authors reviewed and approved the protocol and collected and interpreted the data.

H. Mukae has received funding relevant to the submitted work from Shionogi and grants from Taisho Pharma; lecture fees from Pfizer, MSD, Shionogi, and Taisho Pharma; and advisory fees from Pfizer, MSD, and Shionogi outside the submitted work. H. Yotsuyanagi has received consulting fees from Shionogi, lecture fees from Shionogi and ViiV Healthcare, and travel support from Shionogi outside the submitted work. He serves as an advisory board member of Shionogi and President of the Japanese Society of Infectious Diseases. Y. Doi has received grants from Shionogi and Entasis; consulting fees from Shionogi, Meiji Seika Pharma, Gilead Sciences, GSK, MSD, Chugai, and bioMerieux; and lecture fees from MSD, AstraZeneca, Shionogi, and Teijin Healthcare outside the submitted work and serves as an advisory board member of FujiFilm. H. Sakaguchi, T. Sonoyama, G. Ichihashi, T. Sanaki, K. Baba, Y. Tsuge, and T. Uehara are full-time employees of Shionogi & Co., Ltd., and may have stocks or stock options. N. Ohmagari declares no conflict of interest.

## Author contributions

H. Mukae, H. Yotsuyanagi, N. Ohmagari, Y. Doi, T. Sonoyama, Y. Tsuge, and T. Uehara conceived and designed the experiments and wrote the paper. H. Sakaguchi conceived and designed the experiments, analyzed the data, contributed materials/analysis tools, and wrote the paper. G. Ichihashi conceived and designed the experiments and contributed materials/analysis tools. T. Sanaki conceived and designed the experiments, performed the experiments, analyzed the data, contributed materials/analysis tools, and wrote the paper. K. Baba performed the experiments, analyzed the data, contributed materials/analysis tools, and wrote the paper.

## Extended data

**Extended Data Fig. 1.**
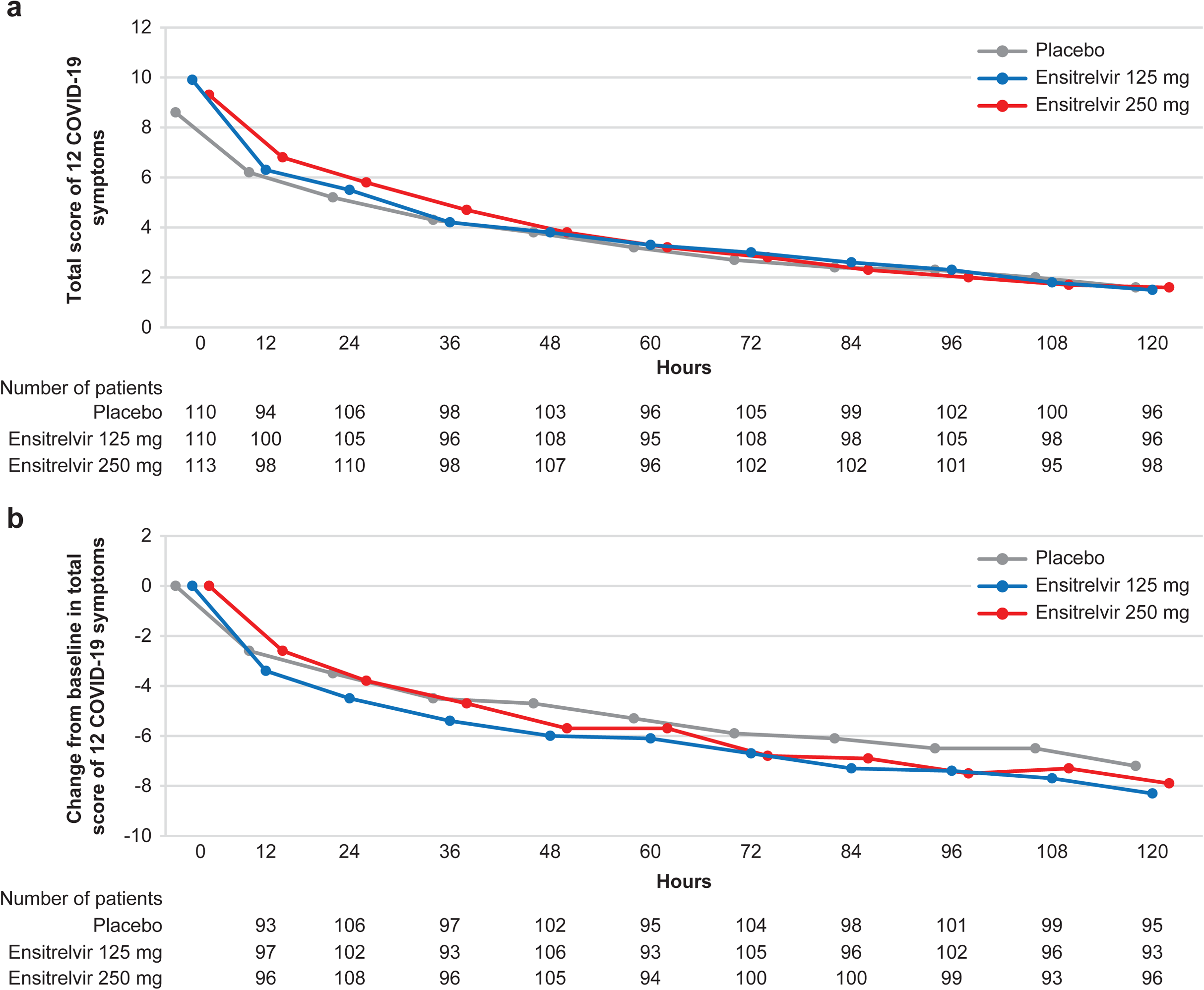
a, Mean absolute values and b, mean change from baseline in the total score of the 12 COVID-19 symptoms (ITT population). COVID-19, coronavirus disease 2019; ITT, intention-to-treat.

**Extended Data Fig. 2.**
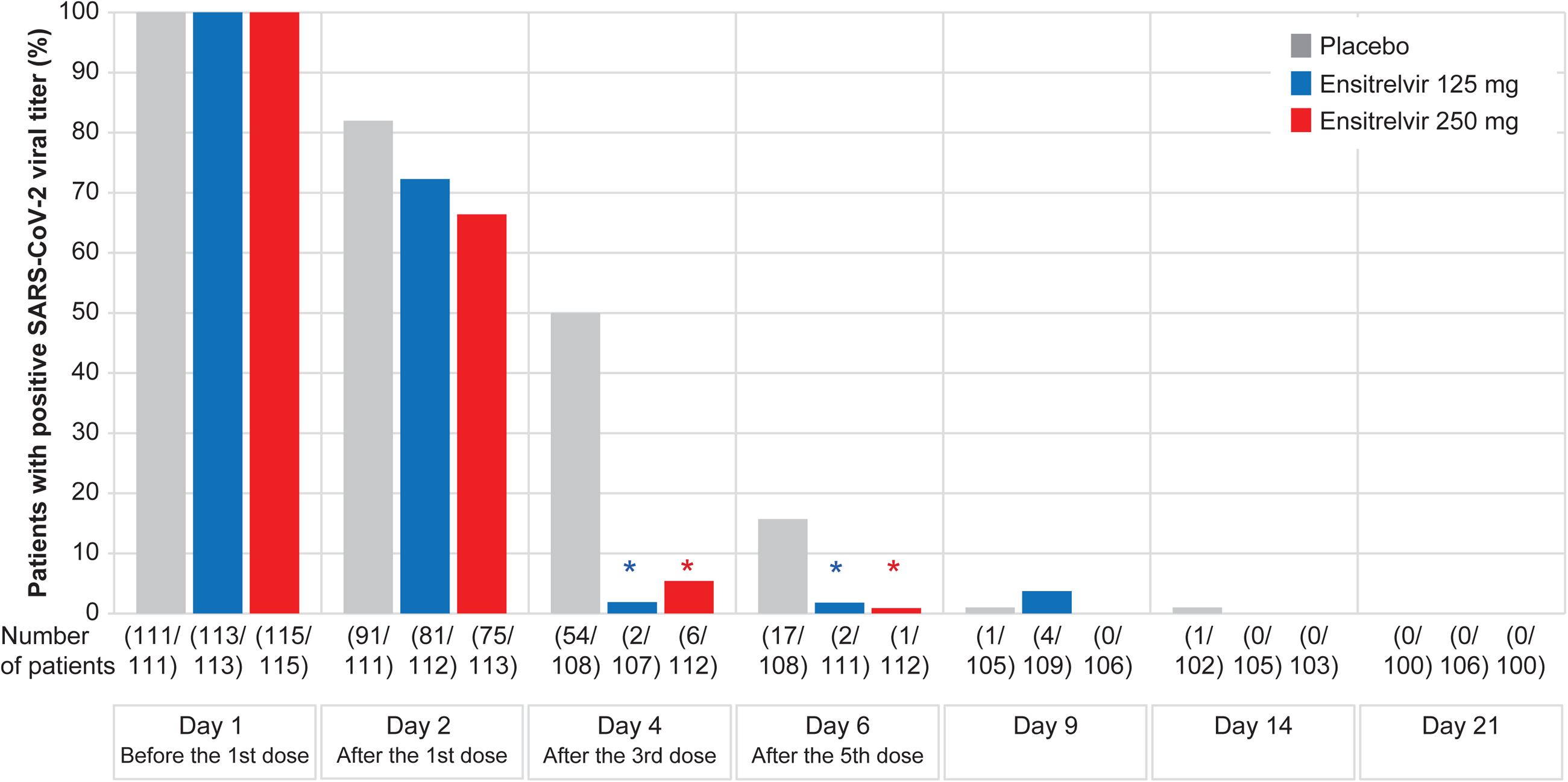
Proportion of patients with a positive SARS-CoV-2 viral titer (ITT population). \**P*<0.05 versus placebo. ITT, intention-to-treat; SARS-CoV-2, severe acute respiratory syndrome coronavirus 2.

**Extended Data Fig. 3.**
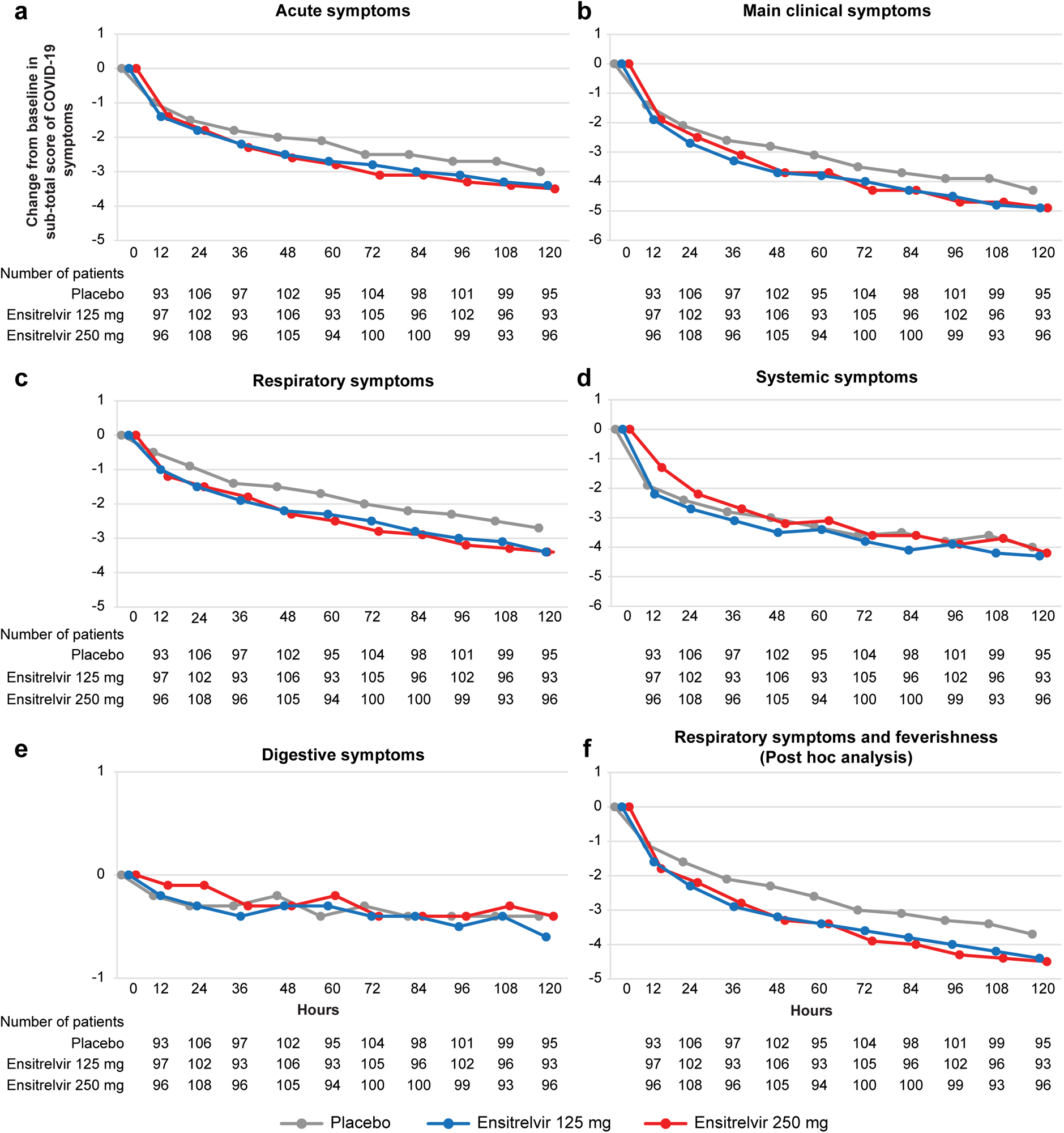
Mean change from baseline in subtotal scores of the 12 COVID-19 symptoms for a, acute symptoms, b, main clinical symptoms, c, respiratory symptoms, d, systemic symptoms, e, digestive symptoms, and f, respiratory symptoms and feverishness (ITT population). COVID-19, coronavirus disease 2019; ITT, intention-to-treat.

**Extended Data Fig. 4.**
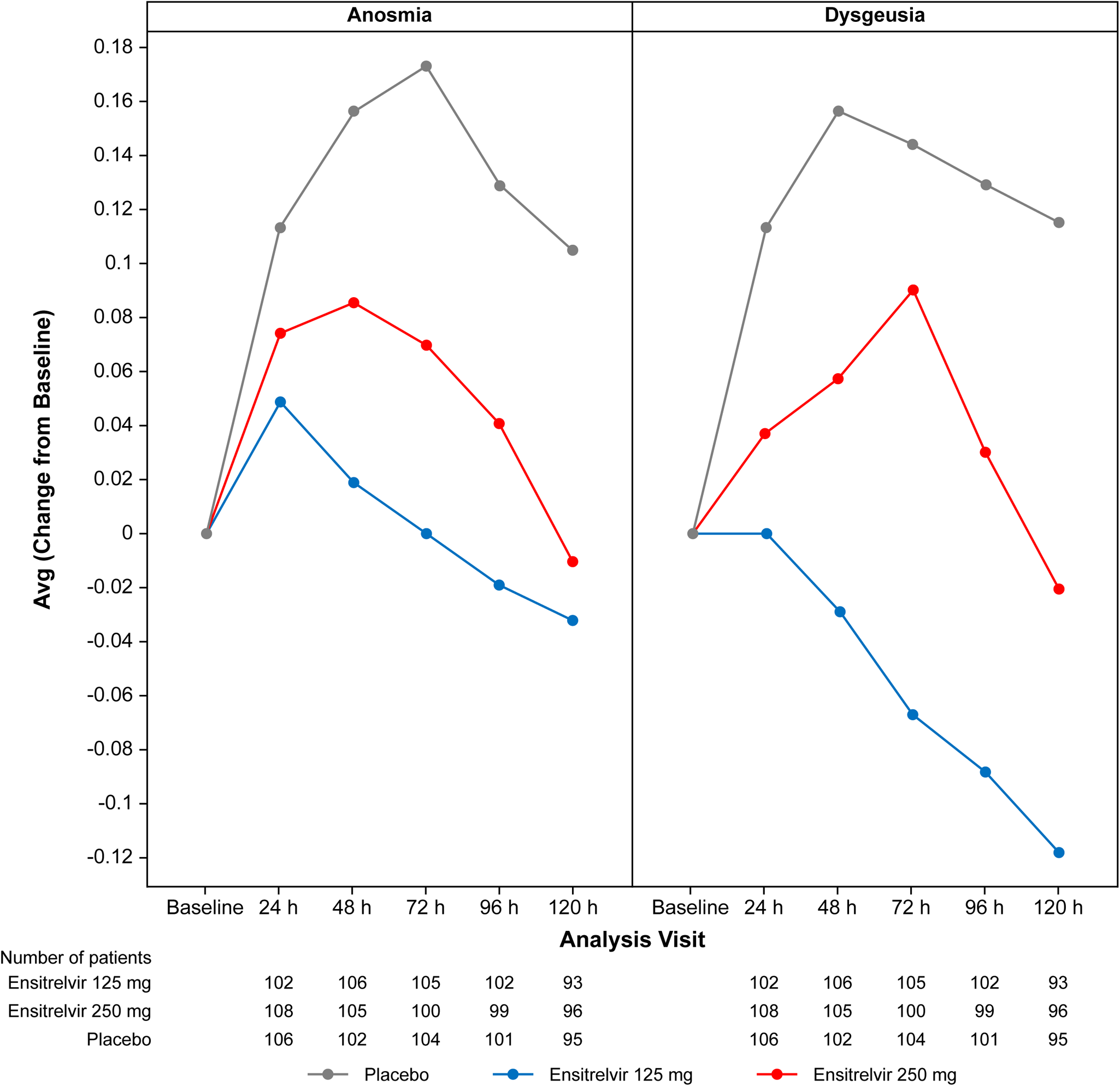
Mean change from baseline in anosmia and dysgeusia scores (ITT population). Avg, average; ITT, intention-to-treat.

**Extended Data Fig. 5.**
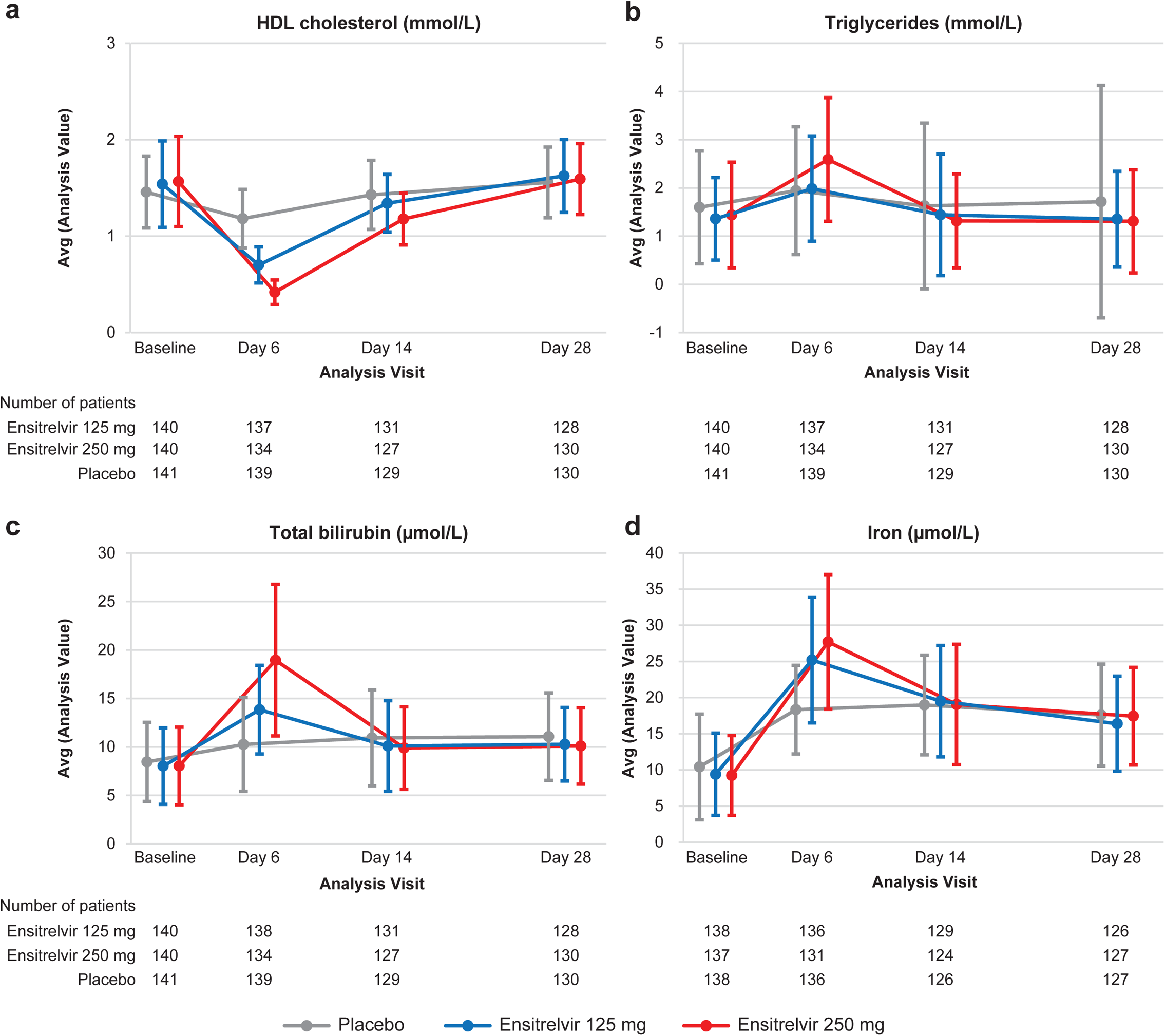
Levels of a, HDL cholesterol, b, triglycerides, c, total bilirubin, and d, iron (safety analysis population). Data are presented as mean±SD. Avg, average; HDL, high-density lipoprotein; SD, standard deviation.

## References

1. Baden, L. R. et al. Efficacy and safety of the mRNA-1273 SARS-CoV-2 vaccine. N. Engl. J. Med. 384, 403–416 (2021).

2. Polack, F. P. et al. Safety and efficacy of the BNT162b2 mRNA Covid-19 vaccine. N. Engl. J. Med. 383, 2603–2615 (2020).

3. Voysey, M. et al. Single-dose administration and the influence of the timing of the booster dose on immunogenicity and efficacy of ChAdOx1 nCoV-19 (AZD1222) vaccine: a pooled analysis of four randomised trials. Lancet 397, 881–891 (2021).

4. Heath, P. T., et al. Safety and efficacy of NVX-CoV2373 Covid-19 vaccine. N. Engl. J. Med. 385, 1172–1183 (2021).

5. Levin, E. G. et al. Waning immune humoral response to BNT162b2 Covid-19 vaccine over 6 months. N. Engl. J. Med. 385, e84 (2021).

6. Bergwerk, M. et al. Covid-19 breakthrough infections in vaccinated health care workers. N. Engl. J. Med. 385, 1474–1484 (2021).

7. Lambrou, A. S. et al. Genomic surveillance for SARS-CoV-2 variants: predominance of the Delta (B.1.617.2) and Omicron (B.1.1.529) variants - United States, June 2021-January 2022. MMWR Morb. Mortal. Wkly. Rep. 71, 206–211 (2022).

8. Ao, D., et al. SARS-CoV-2 Omicron variant: immune escape and vaccine development. MedComm (2020). 3, e126 (2022).

9. Kumar, S. et al. Omicron and Delta variant of SARS-CoV-2: a comparative computational study of spike protein. J. Med. Virol. 94, 1641–1649 (2022).

10. Wolter, N. et al. Early assessment of the clinical severity of the SARS-CoV-2 omicron variant in South Africa: a data linkage study. Lancet 399, 437–446 (2022).

11. Auvigne, V. et al. Severe hospital events following symptomatic infection with Sars-CoV-2 Omicron and Delta variants in France, December 2021 – January 2022: a retrospective, population-based, matched cohort study. eClinicalMedicine doi: 10.1016/j.eclinm.2022.101455 **Error! Hyperlink reference not valid**. (2022).

12. Nyberg, T. et al. Comparative analysis of the risks of hospitalisation and death associated with SARS-CoV-2 omicron (B.1.1.529) and delta (B.1.617.2) variants in England: a cohort study. Lancet 399, 1303–1312 (2022).

13. Jayk Bernal, A., et al. Molnupiravir for oral treatment of Covid-19 in nonhospitalized patients. N. Engl. J. Med. 386, 509–520 (2022).

14. Hammond, J. et al. Oral nirmatrelvir for high-risk, nonhospitalized adults with Covid-19. N. Engl. J. Med. 386, 1397–1408 (2022).

15. Gupta, A. et al. Early treatment for Covid-19 with SARS-CoV-2 neutralizing antibody sotrovimab. N. Engl. J. Med. 385, 1941–1950 (2021).

16. Weinreich, D. M. et al. REGEN-COV antibody combination and outcomes in outpatients with Covid-19. N. Engl. J. Med. 385, e81 (2021).

17. Unoh, Y. et al. Discovery of S-217622, a non-covalent oral SARS-CoV-2 3CL protease inhibitor clinical candidate for treating COVID-19. J. Med. Chem. doi: 10.1021/acs.jmedchem.2c00117 (2022).

18. Uraki, R. et al. Characterization and antiviral susceptibility of SARS-CoV-2 Omicron/BA.2. Nature doi: 10.1038/s41586-022-04856-1 (2022).

19. Sasaki, M., Tabata, K., Kishimoto, M., Itakura, Y., Kobayashi, H., Ariizumi, T., Uemura, K., Toba, S., Kusakabe, S., Maruyama, Y., Iida, S., Nakajima, N., Suzuki, T., Yoshida, S., Nobori, H., Sanaki, T., Kato, T., Shishido, T., Hall, W. W., Orba, Y., Sato, A. & Sawa, H. Oral administration of S-217622, a SARS-CoV-2 main protease inhibitor, decreases viral load and accelerates recovery from clinical aspects of COVID-19. Preprint at bioRxiv https://www.biorxiv.org/content/10.1101/2022.02.14.480338v1.full (2022).

20. Mukae, H., Yotsuyanagi, H., Ohmagari, N., Doi, Y., Imamura, T., Sonoyama, T., Fukuhara, T., Ichihashi, G., Sanaki, T., Baba, K., Takeda, Y., Tsuge, Y. & Uehara, T. A randomized phase 2/3 study of ensitrelvir, a novel oral SARS-CoV-2 3C-like protease inhibitor, in Japanese patients with mild-to-moderate COVID-19 or asymptomatic SARS-CoV-2 infection: results of the phase 2a part. Preprint at medRxiv https://www.medrxiv.org/content/10.1101/2022.05.17.22275027v1 (2022).

21. Fischer, W. A., 2nd, et al. A phase 2a clinical trial of molnupiravir in patients with COVID-19 shows accelerated SARS-CoV-2 RNA clearance and elimination of infectious virus. Sci. Transl. Med. 14, eabl7430 (2022).

22. Mautner, L. et al. Replication kinetics and infectivity of SARS-CoV-2 variants of concern in common cell culture models. Virol. J. 19, 76 (2022).

23. Hay, J. A., Kissler, S. M., Fauver, J. R., Mack, C., Tai, C. G., Samant, R. M., Connelly, S., Anderson, D. J., Khullar, G., MacKay, M., Patel, M., Kelly, S., Manhertz, A., Eiter, I., Salgado, D., Baker, T., Howard, B., Dudley, J. T., Mason, C. E., Ho, D. D., Grubaugh, N. D. & Grad, Y. H. Viral dynamics and duration of PCR positivity of the SARS-CoV-2 Omicron variant. Preprint at medRxiv https://www.medrxiv.org/content/10.1101/2022.01.13.22269257v1 (2022).

24. Singh, R. S. P. et al. Innovative randomized phase I study and dosing regimen selection to accelerate and inform pivotal COVID-19 trial of nirmatrelvir. Clin. Pharmacol. Ther. doi: 10.1002/cpt.2603 (2022).

25. Rodebaugh, T. L. et al. Acute symptoms of mild to moderate COVID-19 are highly heterogeneous across individuals and over time. Open Forum Infect. Dis. 8, ofab090 (2021).

26. Johnson, A. G. et al. COVID-19 incidence and death rates among unvaccinated and fully vaccinated adults with and without booster doses during periods of Delta and Omicron variant emergence - 25 U.S. jurisdictions, April 4-December 25, 2021. MMWR Morb. Mortal. Wkly. Rep. 71, 132–138 (2022).

27. Davies, M-A. et al. Outcomes of laboratory-confirmed SARS-CoV-2 infection in the Omicron-driven fourth wave compared with previous waves in the Western Cape Province, South Africa. Trop. Med. Int. Health. doi: 10.1111/tmi.13752 (2022).

28. Veneti, L. et al. Reduced risk of hospitalisation among reported COVID-19 cases infected with the SARS-CoV-2 Omicron BA.1 variant compared with the Delta variant, Norway, December 2021 to January 2022. Euro. Surveill. 27, 2200077 (2022).

29. Halfmann, P. J. et al. SARS-CoV-2 Omicron virus causes attenuated disease in mice and hamsters. Nature 603, 687–692 (2022).

30. Singanayagam, A. et al. Duration of infectiousness and correlation with RT-PCR cycle threshold values in cases of COVID-19, England, January to May 2020. Euro. Surveill. 25, 2001483 (2020).

31. Blomberg, B. et al. Long COVID in a prospective cohort of home-isolated patients. Nat. Med. 27, 1607–1613 (2021).

32. Lopez-Leon, S. et al. More than 50 long-term effects of COVID-19: a systematic review and meta-analysis. Sci. Rep. 11, 16144 (2021).

33. Pennington, A. F. et al. Risk of clinical severity by age and race/ethnicity among adults hospitalized for COVID-19-United States, March-September 2020. Open Forum Infect. Dis. 8, ofaa638 (2020).

