## Supplementary Tales 1 to 4 for "Efficacy and safety of ensitrelvir in patients with mild-to-moderate COVID-19: the phase 2b part of a randomized, placebo-controlled, phase 2/3 study"

**Supplementary Tables**

**Supplementary Table 1:** List of participating institutions and institutional review boards

| **Institution** | **Institutional review board** |
| --- | --- |
| **Japan** | |
| Kodaira Hospital | Sugiura Clinic Institutional Review Board |
| Swing Nozaki Clinic |  |
| Kasukabe Medical Center |  |
| Japan Community Health care Organization Tokyo Shinjuku Medical Center |  |
| Kouwakai Kouwa Clinic |  |
| Onga Nakama Medical Association Onga Hospital |  |
| Tsuchiura Beryl Clinic |  |
| Edogawa Medicare Hospital |  |
| Kamezawa Clinic |  |
| Ginza Sawada Clinic | Adachi Kyosai Hospital Institutional Review Board |
| Yaguchi Midori Clinic |  |
| Kanagawa Himawari Clinic |  |
| Minna no Tennocho Clinic |  |
| Matsunami General Hospital |  |
| Japan Community Health care Organization Shimonoseki Medical Center |  |
| Irie Clinic |  |
| Fukuoka Shinmizumaki Hospital |  |
| Tashiro Thyroid Clinic |  |
| JA Toride Medical Center | Review Board of Human Rights and Ethics for Clinical Studies Institutional Review Board |
| Moriya Keiyu Hospital |  |
| Japan Community Health care Organization Osaka Minato Central Hospital |  |
| Kinki Central Hospital of the Mutual Aid Association of Public School Teachers |  |
| Medical Corporation Kouhoukai Takagi Hospital |  |
| Fujimaki Ent Clinic | Yoga Allergy Clinic Institutional Review Board |
| Takinogawa Hospital |  |
| Kaiseikai Kita Shin Yokohama Internal Medicine Clinic |  |
| Nakahama Clinic |  |
| Lee Clinic |  |
| National Hospital Organization Kanazawa Medical Center | National Hospital Organization Kanazawa Medical Center Institutional Review Board |
| Koga General Hospital | Tokushukai Group Institutional Review Board |
| Kamagaya General Hospital |  |
| Nozaki Tokushukai Hospital |  |
| Ikoma City Hospital |  |
| Showa University Hospital | Showa University Hospital Institutional Review Board |
| Showa University East Hospital |  |
| Yokohama Municipal Citizen's Hospital | Yokohama Municipal Citizen's Hospital Institutional Review Board |
| Aso Iizuka Hospital | Aso Iizuka Hospital Institutional Review Board |
| Red Cross Kyoto Daini Hospital | Japanese Red Cross Society Japanese Red Cross Kyoto Daini Hospital Institutional Review Board |
| University Hospital Kyoto Prefectural University of Medicine | University Hospital Kyoto Prefectural University of Medicine Institutional Review Board |
| Tokyo Metropolitan Health and Hospitals Corporation Toshima Hospital | Tokyo Metropolitan Health and Hospitals Toshima Hospital Institutional Review Board |
| IUHW Narita Hospital | International University of Health and Welfare and Institutional Review Board |
| Center Hospital of the National Center for Global Health and Medicine | Center Hospital of National Center for Global Health and Medicine IRB |
| Hokkaido University Hospital | Hokkaido University Hospital Institutional Review Board |
| Sapporo Medical University Hospital | Sapporo Medical University Hospital Institutional Review Board |
| Denenchofu Family Clinic | Kobori Central Clinical Research Ethics Committee |
| Osaka Rosai Hospital | Osaka Rosai Hospital Institutional Review Board |
| Kojunkai Daido Medicine | Daido Hospital Institutional Review Board |
| Rinku General Medical Center | Rinku General Medical Center IRB |
| University of Tsukuba Hospital | Institutional Review Board, University of Tsukuba Hospital |
| Nagasaki University Hospital | Nagasaki University Hospital Institutional Review Board |
| Tokyo Medical University Hachioji Medical Center | Tokyo Medical University Hachioji Medical Center Institutional Review Board |
| IMSUT Hospital, The Institute of Medical Science, The University of Tokyo | IMSUT Hospital, The Institute of Medical Science, The University of Tokyo Institutional Review Board |
| Fujita Health University Okazaki Medical Center | The Central Institutional Review Board for the Fujita Health University Hospitals |
| National Hospital Organization Yokohama Medical Center | National Hospital Organization Yokohama Medical Center Institutional Review Board |
| Okinawa National Hospital | Okinawa National Hospital Clinical Trial Review Committee |
| National Hospital Organization Kobe Medical Center | National Hospital Organization Kobe Medical Center Institutional Review Board |
| National Hospital Organization Himeji Medical Center | National Hospital Organization Himeji Medical Center Institutional Review Board |
| National Hospital Organization Fukuoka National Hospital | National Hospital Organization Fukuoka National Hospital Institutional Review Board |
| National Hospital Organization Chiba Medical Center | National Hospital Organization Chiba Medical Center Institutional Review Board |
| Japan Community Health Care Organization Hokkaido Hospital | Review Board of Japan Community Health care Organization Hokkaido Hospital |
| Hamamatsu Medical Center | Hamamatsu Medical Center Institutional Review Board |
| Nagoya City University East Medical Center | NCU East/West Medical Center Institutional Review Board |
| **South Korea** | |
| Gachon University Gil Medical Center | Gachon University Gil Medical Center Institutional Review Board |
| The Catholic Univ. of Korea Bucheon St. Mary’s Hospital | The Catholic Medical Center Central Institutional Review Board |
| The Catholic Univ. of Korea Eunpyeong St. Mary’s Hospital | The Catholic Medical Center Central Institutional Review Board |
| Gangnam Severance Hospital | Gangnam Severance Hospital Institutional Review Board |
| KyungHee University Hospital at Gangdong | Kyung Hee University Hospital at Gangdong, Institutional Review Board |
| Kyungpook National University Hospital | Kyungpook National University Hospital IRB |
| KyungHee University Medical Center | Kyung Hee University Hospital, Institutional Review Board |
| Keimyung University Daegu Dongsan Hospital | Keimyung University Daegu dongsan Medical Center Institutional Review Board |
| Korea University Anam Hospital | Korea University Hospital IRB |
| Dongguk University Ilsan Hospital | Dogguk University Ilsan Hospital Institutional Review Board |
| Bupyeong Serim Hospital | IRB of Bupyeong Serim Hospital |
| Yongin Severance Hospital | YONGIN SEVERANCE  HOSPITAL Institutional Review Board |
| Uijeongbu Eulji Medical Center, Eulji University | Uijeongbu Eulji Medical Center, Eulji University Institutional Review Board |
| Ewha Womans University Mokdong Hospital | Ewha Womans University Mokdong Hospital Institutional Review Board |
| Inje University Ilsan Paik Hospital | Inje University Ilsan Paik Hospital IRB |
| Incheon Medical Center | Incheon Medical Center Institutional Review Board |
| Inha University Hospital | Inha University Hospital IRB |
| Chung-Ang University Health Care System Hyundae Hospital | Chung-Ang University Healthcare System Hyundae Hospital Institutional Review Board |
| Chung-Ang University Hospital | Chung-Ang University Hospital Institutional Review Board |
| VHS Medical Center | VHS Medical Center Institutional Review Board |
| Kyungpook National University Chilgok Hospital | Kyungpook National University Chilgok Hospital Institutional Review Board |
| SMG-SNU Boramae Medical Center | Seoul Metropolitan Government Seoul National University Boramae Medical Center Institutional Review Board |
| Incheon Sejong Hospital | Incheon Sejong Hospital Institutional Review Board |
| Myongji Hospital | Myongji Hospital Institutional Review Board |
| Chungnam National University Sejong Hospital | Chungnam National University Sejong Hospital Institutional Review Board |

**Supplementary Table 2:** Questionnaire and symptom subscales for the 14 COVID-19 symptoms

| **Questionnaire item** | **Symptom subscale** | | | | | |
| --- | --- | --- | --- | --- | --- | --- |
|  | **Acute symptoms** | **Main clinical symptoms** | **Respiratory symptoms** | **Systemic symptoms** | **Digestive symptoms** | **Respiratory symptoms and feverishness** |
| 1. Stuffy or runny nose*^a^* |  | ● | ● |  |  | ● |
| 2. Sore throat*^a^* | ● | ● | ● |  |  | ● |
| 3. Shortness of breath (difficulty breathing)*^a^* |  |  | ● |  |  | ● |
| 4. Cough*^a^* | ● | ● | ● |  |  | ● |
| 5. Low energy or tiredness*^a^* |  |  |  | ● |  |  |
| 6. Muscle or body aches*^a^* |  |  |  | ● |  |  |
| 7. Headache*^a^* |  |  |  | ● |  |  |
| 8. Chills or shivering*^a^* |  | ● |  | ● |  |  |
| 9. Feeling hot or feverish*^a^* | ● | ● |  | ● |  | ● |
| 10. Nausea (feeling like you want to throw up)*^a^* |  |  |  |  | ● |  |
| 11. Vomiting (throwing up) |  |  |  |  | ● |  |
| 12. Diarrhea (loose or watery stools) |  |  |  |  | ● |  |
| 13. Rate your sense of smell in the last 24 hours*^b^* |  |  |  |  |  |  |
| 14. Rate your sense of taste in the last 24 hours*^b^* |  |  |  |  |  |  |

For items 1 to 12, each symptom was rated on a 4-point scale (None=0, Mild=1, Moderate=2, or Severe=3). For items 13 and 14, the sense of smell/taste was rated on a 3-point scale (Same as usual=0, Less than usual=1, or No sense of smell/taste=2).

Time to improvement of COVID-19 symptoms was defined as the time from the study intervention initiation to the time when all symptoms met the following criteria:

- Symptoms that were present prior to COVID-19 onset and considered by the patient to have worsened at baseline: severe symptoms improving to moderate or better or moderate symptoms improving to mild or better, persisting for 24 hours.
- Symptoms that were present prior to COVID-19 onset and considered by the patient not to have worsened at baseline: severe symptoms remaining severe or improving or moderate symptoms remaining moderate or improving, persisting for 24 hours.
- Symptoms that were not present prior to COVID-19 onset but occurred after baseline: mild or better condition persisting for 24 hours.

*^a^*Severity of the symptom at its worst over the last 24 hours.

*^b^*Not used to calculate the total score of the 12 COVID-19 symptoms.

COVID-19, coronavirus disease 2019.

**Supplementary Table 3:** Change from baseline in SARS-CoV-2 viral titer (log_10_ TCID_50_/mL) on day 4 stratified by COVID-19 vaccination history and time from onset to randomization (ITT population)

| **Statistics** | **Ensitrelvir 125 mg**  **(*N=*114)** | **Ensitrelvir 250 mg**  **(*N=*116)** | **Placebo**  **(*N=*111)** |
| --- | --- | --- | --- |
| COVID-19 vaccination (Yes) |  |  |  |
| *N* | 89 | 93 | 93 |
| Mean (SD) change from baseline | -1.66 (0.85) | -1.44 (0.83) | -1.05 (1.00) |
| LS mean (SE) change from baseline assessed by ANCOVA | -1.51 (0.04) | -1.50 (0.04) | -1.13 (0.04) |
| LS mean (SE) difference in change from baseline versus placebo | -0.37 (0.05) | -0.37 (0.05) |  |
| 95% CI | -0.47 to -0.27 | -0.47 to -0.27 |  |
| *P*-value | <0.0001 | <0.0001 |  |
| COVID-19 vaccination (No) |  |  |  |
| *N* | 17 | 19 | 14 |
| Mean (SD) change from baseline | -1.81 (0.81) | -1.34 (0.84) | -1.11 (0.96) |
| LS mean (SE) change from baseline assessed by ANCOVA | -1.62 (0.12) | -1.59 (0.11) | -0.98 (0.13) |
| LS mean (SE) difference in change from baseline versus placebo | -0.64 (0.17) | -0.61 (0.17) |  |
| 95% CI | -0.98 to -0.29 | -0.96 to -0.27 |  |
| *P*-value | 0.0006 | 0.0008 |  |
| Time from onset to randomization (<72 hours) |  |  |  |
| *N* | 52 | 52 | 53 |
| Mean (SD) change from baseline | -1.71 (0.79) | -1.64 (0.80) | -1.02 (1.28) |
| LS mean (SE) change from baseline assessed by ANCOVA | -1.64 (0.07) | -1.64 (0.07) | -0.93 (0.07) |
| LS mean (SE) difference in change from baseline versus placebo | -0.71 (0.09) | -0.71 (0.09) |  |
| 95% CI | -0.89 to -0.54 | -0.88 to -0.54 |  |
| *P*-value | <0.0001 | <0.0001 |  |
| Time from onset to randomization (≥72 hours) |  |  |  |
| *N* | 54 | 60 | 54 |
| Mean (SD) change from baseline | -1.67 (0.89) | -1.24 (0.81) | -1.09 (0.59) |
| LS mean (SE) change from baseline assessed by ANCOVA | -1.35 (0.03) | -1.35 (0.03) | -1.22 (0.03) |
| LS mean (SE) difference in change from baseline versus placebo | -0.13 (0.04) | -0.13 (0.04) |  |
| 95% CI | -0.20 to -0.05 | -0.20 to -0.05 |  |
| *P*-value | 0.0013 | 0.0007 |  |

ANCOVA, analysis of covariance; CI, confidence interval; COVID-19, coronavirus disease 2019; ITT, intention-to-treat; LS, least squares; SARS-CoV-2, severe acute respiratory syndrome coronavirus 2; SD, standard deviation; SE, standard error; TCID_50_, 50% tissue-culture infectious dose.

**Supplementary Table 4:** Time-weighted average change from baseline up to 120 hours after initial drug administration in the subtotal scores of 12 COVID-19 symptoms (ITT population)

| **Statistics** | **Ensitrelvir 125 mg**  **(*N=*114)** | **Ensitrelvir 250 mg**  **(*N=*116)** | **Placebo**  **(*N=*111)** |
| --- | --- | --- | --- |
| Acute symptoms |  |  |  |
| *N* | 107 | 108 | 109 |
| Mean (SD) change from baseline | -2.52 (1.34) | -2.63 (1.34) | -2.09 (1.31) |
| LS mean (SE) change from baseline assessed by ANCOVA | -2.36 (0.10) | -2.50 (0.10) | -2.16 (0.10) |
| LS mean (SE) difference in change from baseline versus placebo | -0.20 (0.12) | -0.33 (0.12) | --- |
| 95% CI | -0.44 to 0.04 | -0.58 to -0.09 | --- |
| *P*-value | 0.1080 | 0.0070 | --- |
| Main clinical symptoms |  |  |  |
| *N* | 109 | 111 | 110 |
| Mean (SD) change from baseline | -3.56 (2.01) | -3.54 (2.00) | -2.94 (1.94) |
| LS mean (SE) change from baseline assessed by ANCOVA | -3.34 (0.14) | -3.42 (0.14) | -2.99 (0.14) |
| LS mean (SE) difference in change from baseline versus placebo | -0.34 (0.17) | -0.42 (0.17) | --- |
| 95% CI | -0.68 to 0.00 | -0.76 to -0.08 | --- |
| *P*-value | 0.0504 | 0.0149 | --- |
| Respiratory symptoms |  |  |  |
| *N* | 106 | 111 | 109 |
| Mean (SD) change from baseline | -2.28 (1.54) | -2.33 (1.58) | -1.67 (1.44) |
| LS mean (SE) change from baseline assessed by ANCOVA | -2.09 (0.12) | -2.16 (0.12) | -1.72 (0.13) |
| LS mean (SE) difference in change from baseline versus placebo | -0.37 (0.15) | -0.44 (0.15) | --- |
| 95% CI | -0.67 to -0.07 | -0.74 to -0.15 | --- |
| *P*-value | 0.0153 | 0.0033 | --- |
| Systemic symptoms |  |  |  |
| *n* | 100 | 104 | 98 |
| Mean (SD) change from baseline | -3.71(2.86) | -3.19 (2.54) | -3.35 (2.28) |
| LS mean (SE) change from baseline assessed by ANCOVA | -3.31 (0.14) | -3.16 (0.14) | -3.46 (0.14) |
| LS mean (SE) difference in change from baseline versus placebo | 0.15 (0.18) | 0.30 (0.17) | --- |
| 95% CI | -0.20 to 0.49 | -0.04 to 0.64 | --- |
| *P*-value | 0.4052 | 0.0843 | --- |
| Digestive symptoms |  |  |  |
| *n* | 35 | 35 | 32 |
| Mean (SD) change from baseline | -1.39 (1.18) | -1.13 (1.37) | -1.27 (0.86) |
| LS mean (SE) change from baseline assessed by ANCOVA | -1.27 (0.14) | -1.08 (0.14) | -1.32 (0.14) |
| LS mean (SE) difference in change from baseline versus placebo | 0.06 (0.18) | 0.25 (0.18) | --- |
| 95% CI | -0.30 to 0.42 | -0.11 to 0.61 | --- |
| *P*-value | 0.7519 | 0.1769 | --- |
| Respiratory symptoms and feverishness (post hoc analysis) |  |  |  |
| *n* | 108 | 111 | 109 |
| Mean (SD) change from baseline | -3.17 (1.79) | -3.26 (1.81) | -2.49 (1.66) |
| LS mean (SE) change from baseline assessed by ANCOVA | -2.97 (0.13) | -3.04 (0.13) | -2.56 (0.14) |
| LS mean (SE) difference in change from baseline versus placebo | -0.40 (0.17) | -0.48 (0.16) | --- |
| 95% CI | -0.73 to -0.07 | -0.80 to -0.15 | --- |
| *P*-value | 0.0164 | 0.0039 | --- |

ANCOVA, analysis of covariance; CI, confidence interval; COVID-19, coronavirus disease 2019; ITT, intention-to-treat; LS, least squares; SD, standard deviation; SE, standard error.
